# Sex-related signatures in the plasma proteome not attributable to XX and XY chromosomes and their impact on health and disease

**DOI:** 10.64898/2026.10.19.26365875

**Authors:** Essi Viippola, Tuomo Hartonen, Meihui Zhu, Zhijian Yang, Ryuya Edahiro, Tomoko Nakanishi, Ao Lan, Masahiro Kanai, Nhi N. L. Nguyen, Tatiana Cajuso, Daniela Fusco, Chiara Caime, Kristina Zguro, Sebastian May-Wilson, M. Austin Argentieri, Gabin Drouard, Lisa Eick, Matteo Ferro, Aoxing Liu, Rodosthenis Rodosthenous, FinnGen, Biobank Japan, Japan COVID-19 Task Force, Jarno Honkanen, Jukka Partanen, Miina Ollikainen, Jaakko Kaprio, Ho Namkoong, Koichi Fukunaga, Koichi Matsuda, Jerome I. Rotter, Stephen S. Rich, Kent D. Taylor, Robert E. Gerszten, Clary Clish, Peter Y. Liu, Alexis C. Wood, Craig Johnson, Ada S. Cheung, Tamar Sofer, Mark Daly, Taru Tukiainen, Boris Novakovic, Yukinori Okada, Xia Shen, Andrea Ganna

**Affiliations:** Institute for Molecular Medicine Finland (FIMM), HiLIFE, University of Helsinki, Helsinki, Finland; Program in Medical and Population Genetics, Broad Institute of MIT and Harvard, Cambridge, MA, USA; Analytic and Translational Genetics Unit, Department of Medicine, Massachusetts General Hospital, Boston, MA, USA; Department of Statistical Genetics, Osaka University Graduate School of Medicine, Suita, Japan; Department of Respiratory Medicine and Clinical Immunology, Osaka University Graduate School of Medicine, Suita, Japan; Laboratory for Systems Genetics, RIKEN Center for Integrative Medical Sciences, Yokohama, Japan; Department of Genome Informatics, Graduate School of Medicine, University of Tokyo, Tokyo, Japan; Precision Healthcare University Research Institute, Queen Mary University of London, London, UK; Center for Intelligent Medicine Research, Greater Bay Area Institute of Precision Medicine (Guangzhou), Fudan University, Guangzhou, China; State Key Laboratory of Genetic Engineering, Center for Evolutionary Biology, School of Life Sciences, Fudan University, Shanghai, China; Stanley Center for Psychiatric Research, Broad Institute of MIT and Harvard, Cambridge, MA, USA; Department of Neurosciences “Rita Levi Montalcini”, University of Turin, Italy; Department of Medicine and Surgery, University of Milano-Bicocca, Monza, Italy; Division of Gastroenterology, Center for Autoimmune Liver Diseases, European Reference Network on Hepatological Diseases (ERN RARE-LIVER), Fondazione IRCCS San Gerardo dei Tintori, Monza, Italy; Finnish Red Cross Blood Service, Helsinki, Finland; Minerva Foundation Institute for Medical Research, Helsinki, Finland; Department of Infectious Diseases, Keio University School of Medicine, Tokyo, Japan; Division of Pulmonary Medicine, Department of Medicine, Keio University School of Medicine, Tokyo, Japan; Laboratory of Clinical Genome Sequencing, Department of Computational Biology and Medical Sciences, Graduate School of Frontier Sciences, University of Tokyo, Tokyo, Japan; Institute of Medical Science, The University of Tokyo, Tokyo, Japan; The Institute for Translational Genomics and Population Sciences, Department of Pediatrics, The Lundquist Institute for Biomedical Innovation at Harbor-UCLA Medical Center, Torrance, CA, USA; Center for Public Health Genomics, Department of Public Health Sciences, University of Virginia, Charlottesville, VA, USA; Department of Medicine, Beth Israel Deaconess Medical Center, Boston, MA, USA; Broad Institute of MIT and Harvard, Cambridge, MA, USA; Division of Genomic Outcomes, Departments of Pediatrics and Medicine, Harbor-UCLA Medical Center, Torrance, CA, USA; Division of Endocrinology, Department of Medicine, The Lundquist Institute at Harbor-UCLA Medical Center, Torrance, CA, USA; US Department of Agriculture/Agricultural Research Service Children’s Nutrition Research Center, Children’s Nutrition Research Center, Baylor College of Medicine, Houston, TX, USA; University of Washington, Seattle, WA, USA; Department of Medicine, Harvard Medical School, Boston, MA, USA; Department of Biostatistics, Harvard T.H. Chan School of Public Health, Boston, MA, USA; Division of Cardiology, Department of Medicine, Beth Israel Deaconess Medical Center, Boston, MA, USA; Institute of Clinical Medicine, School of Medicine, Faculty of Health Sciences, University of Eastern Finland, Kuopio, Finland; Department of Medical Epidemiology and Biostatistics, Karolinska Institutet, Stockholm, Sweden; Faculty of Medicine, University of Helsinki, Helsinki, Finland; Molecular Immunity, Infection, Immunity and Global Health Theme, Murdoch Children’s Research Institute, Royal Children’s Hospital, Parkville, Victoria, Australia; Department of Paediatrics, The University of Melbourne, Parkville, Victoria, Australia; Department of Medicine (Austin Health), The University of Melbourne, Heidelberg, Victoria, Australia; Department of Endocrinology, Austin Health, Heidelberg, Victoria, Australia

## Abstract

Sex differences in human health are typically modeled as binary comparisons between males (XY) and females (XX). However, sex-linked molecular traits vary continuously and are shaped by environmental factors. Here, we introduce the Proteomic Sex Index (PSI), a continuous, plasma proteomics–based measure that quantifies the degree to which an individual’s proteome aligns with their chromosomal sex, with higher PSI values indicating profiles less typical for their sex. Leveraging data from 51,310 individuals from six demographically heterogeneous cohorts, we show that PSI captures broad, biologically meaningful variation in sex-enriched traits and diseases. Individuals with proteomic profiles shifted away from their chromosomal sex exhibited trait values less characteristic of that sex. For example, males with higher PSI were shorter, had lower grip strength, and showed reduced severity of COVID-19 (OR=0.59, *p*=3.0×10^−7^). Analyses of individuals undergoing gender-affirming hormone therapy showed that, although PSI correlated with circulating sex hormones, it also captured additional variation independent of hormone levels. PSI is partially heritable and genetically influenced. Individuals with Klinefelter syndrome (XXY) had markedly elevated PSI (OR=13.7, *p*=1.1×10^−13^), and genome-wide association studies of PSI identified sex-biased cis-pQTLs with evidence of causal links to sex-specific disease risk. PSI provides a nuanced, individualized measure of sex-related biology that enables the study of within-sex molecular variation and its links to health and disease.

## Main

Sex characteristics play a crucial role in human biology. Many diseases and traits exhibit sex differences ^1^, and understanding these differences is a major scientific and societal priority. Males and females differ at the molecular level; for example, at the transcriptomic level, more than 30% of genes are differentially expressed between sexes in at least one tissue ^2^. While binary comparisons, typically based on chromosomal sex (i.e., XX or XY chromosomes; **Box 1**), provide a starting point, they do not fully capture the complexity of sex differences in biology. Binary comparisons capture “average” effects between sexes, but substantial variability exists within each chromosomal sex. While sex-specific average expressions of sex-biased genes differ ^2^, the ranges of expression values still largely overlap between sexes. Multiple factors can contribute to this variability. Genetic effects, such as sex chromosome aneuploidies, can lead to physiological traits that diverge from XX or XY patterns ^3^. Environmental and societal factors, including gender-related exposures and diagnostic biases, also play a role ^1,4^. Additionally, biological events such as menopause introduce time-dependent changes to sex-related molecular traits. Together, these influences result in substantial heterogeneity in sex-associated molecular signatures even among individuals sharing the same sex chromosome makeup. A helpful, albeit imperfect, analogy ^5^ is biological aging: although chronological age is fixed, individuals age at different rates, and deviation from the average trajectory is determined by a complex interplay of genetic and environmental factors.

In this study, sex is defined based on chromosomal information: individuals with XX chromosomes are referred to as females, and individuals with XY chromosomes as males. Analyses considering other sex chromosome configurations are highlighted separately. Throughout the text, the terms “male” and “female” refer strictly to chromosomal sex and do not encompass other definitions of sex, such as sex assigned at birth, and do not refer to gender identity, although we recognize the importance of these factors in explaining health outcomes independently of chromosomal sex. Even though we do not have information about individuals’ gender identity, it is likely that most individuals included in the study are cisgender. In addition, one component of the study includes transgender and non-binary individuals undergoing gender-affirming hormone therapy. Accompanying this paper, we provide a general audience summary (see **Supplementary Materials**) communicating the main motivations, findings, and their interpretation.

In this study, we focus on quantifying and characterizing sex-related molecular variation that makes some individuals more similar to—or different from—their peers of the same chromosomal sex. We further investigate whether this variation can help explain differences in traits and disease risk between sexes that are not attributable solely to XX or XY chromosomal differences.

To address these questions, we leverage broad-capture plasma proteomics. Plasma proteomics offers key advantages: it captures proteins originating from tissues throughout the body, integrates both genetic and environmental influences, and provides a systemic view of organ-specific biology through a single, accessible medium ^6^. Plasma proteins reflect the functional outcomes of gene regulation, and proteomic profiles have demonstrated strong predictive power across a broad range of diseases ^7–10^. These properties make plasma proteomics a powerful platform for studying biological variability related to sex.

Substantial sex differences in the plasma proteome have been reported across the life course, including in adolescents ^11^ and adults ^12–14^. Building on these results, we develop the Proteomic Sex Index (PSI), a machine learning–derived measure that quantifies how closely an individual’s plasma proteome aligns with proteomic patterns common among individuals sharing the same sex chromosome configuration. PSI provides a continuous, dynamic representation of proteomic sex-related variation, enabling systematic investigations into its relationships with sex-linked traits, genetic factors, and disease risk.

## Results

### Most plasma proteins are sex-biased and replicate in studies with different demographic and ancestry characteristics

We compared the relative abundance of 2,919 proteins measured in plasma from N=42,979 participants in the UK Biobank ^15,16^ (54.1 % females (XX sex chromosomes), median age 58; **Supplementary Table 1**). A vast majority (80 %) of proteins demonstrated significant age-adjusted sex-specific expression (Bonferroni-corrected p-value threshold 2×10^−5^), and out of those, 273 proteins had a fold change > 1.5, underscoring the widespread nature of sex differences at the proteomic level. Among these highly sex-biased proteins, 60 % had higher levels in males (XY sex chromosomes; male-biased proteins), while the remainder were female-biased (**Supplementary Table 2**).

To confirm the robustness of sex bias effects across proteins, we repeated the analyses in four additional studies: FinnGen ^17^ (N=1,199), Multi-Ethnic Study of Atherosclerosis ^18^ (MESA; N=2,851), Biobank Japan ^19^ (BBJ; N=2,870), and Japan COVID-19 Task Force ^20^ (JCTF; N=1,372). Compared to the UK Biobank, FinnGen is considerably younger, MESA consists of almost 60 % non-European ancestries, and the Japanese cohorts, BBJ and JCTF, are predominantly of East Asian ancestry,consist of around 75 % males, and have a higher disease burden due to disease-based recruitment. Despite differences in demographics and study designs (**Supplementary Table 1**), the magnitude of sex bias effects showed good concordance with the UK Biobank (rank correlation between coefficients=0.64 to 0.82; **Supplementary Table 3**). This replication analysis shows that sex bias of plasma protein expression is relatively stable across cohorts of varying genetic ancestries and age distributions.

### Development of the Proteomic Sex Index (PSI) to capture within-sex variability

Despite the widespread sex differences among proteins, no protein completely separates males and females (**Supplementary Table 2**). Even for proteins with the largest sex differences, which reflect sex-specific biological function and are often expressed by organs present only in males or females, we observe large variability *within* the chromosomal sexes and overlap *between* sexes (**Supplementary Fig. 1**). For example, Epididymal Protein 3B (EDDM3B) has a possible function in sperm maturation, is almost exclusively expressed in the epididymis, and is strongly male-biased (i.e. it has higher expression in males than females). Nonetheless, in the UK Biobank, there were 215 females (out of 23,242, 0.93 %) whose EDDM3B levels were higher than 50 % of males. Similarly, XG is one of the strongest female-biased proteins, but we still observe 958 males (out of 19,736, 4.85%) with protein levels higher than the female median.

Motivated by these observations, we sought to quantify broader sex-related proteomics variation within each chromosomal sex. We trained an eXtreme Gradient Boosting ^21^ (XGBoost) machine learning model that uses protein expression to predict chromosomal sex (XX *vs.* XY) in the UK Biobank training set (N=9,128; 20 % of the UK Biobank individuals with Olink proteomics data; **Figure 1**). Importantly, we age-matched males and females in the training set to avoid the model from learning any age differences between males and females. We also removed from the training set any individuals with discordant chromosomal and self-reported sex, as well as individuals with sex chromosome aneuploidies. We performed feature selection based on SHapley Additive exPlanations ^22^ (SHAP) values using the Stochastic Gradient Boosting ensembles approach ^23^. This means we trained 1,000 different XGBoost models and selected only the most robustly important proteins as features in the final prediction model (see **Methods** for details). For each individual, we obtained the XGBoost-predicted male probability (XY sex chromosomes), which we refer to as the *proteomics-predicted sex*, and computed the difference between the proteomics-predicted sex and the observed chromosomal sex (coded as 0 for females and 1 for males). We define the *Proteomic Sex Index (PSI)* as its inverse rank-normalized transformation within each sex. PSI captures within-sex variability in the sex-linked proteome. Higher values indicate higher similarity of the sex-associated plasma proteome with patterns commonly observed in individuals with different chromosomal sex.

**Fig. 1.**
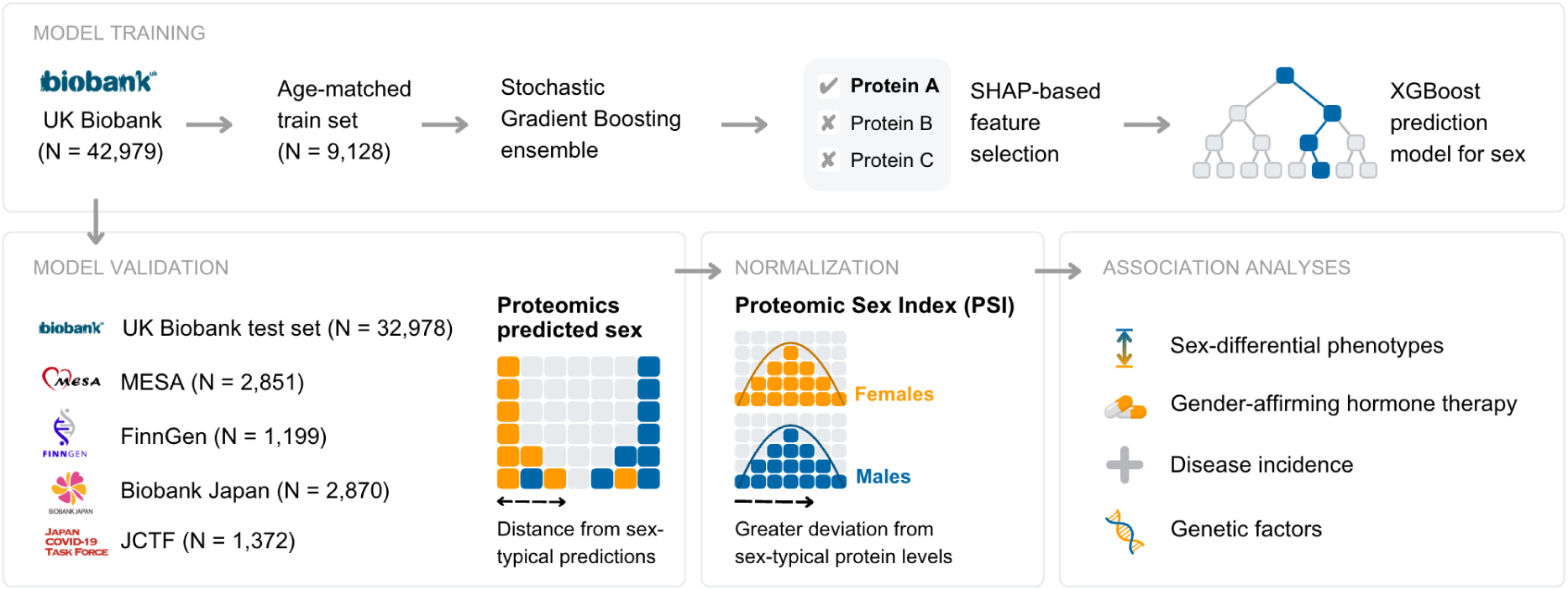
Proteomics-based prediction model for chromosomal sex. Schematic overview of the training and validation of a continuous proteomics-based predictor of chromosomal sex (XX or XY sex chromosomes). An XGBoost model was trained on 20 % of the UK Biobank proteomics dataset (N = 9,128). After SHAP value-based feature selection, the final model included 31 protein features. Model performance was evaluated on an unseen UK Biobank test set and further validated in four external cohorts. For each individual, we calculated the difference between the XGBoost-predicted probability and the observed chromosomal sex and applied inverse rank-normalization within each sex to derive the Proteomic Sex Index (PSI). We then assessed associations between PSI and sex-differential phenotypes, gender-affirming hormone therapy, disease incidence, and genetic factors.

The final model includes 31 proteins, the majority of which (71 %) were male-biased in univariate analyses (**Figure 2a**). These proteins span a broad range of biological functions. The most predictive proteins—ranked by mean absolute SHAP values (**Figure 2b**; **Supplementary Table 4**)—include KLK3, INSL3, and ACRV1, all of which are integral to reproductive biology, and specifically, male reproductive function. Other proteins selected by the model are involved in muscle development and tissue remodeling (e.g., SOST, CDH15, ACAN), cardiometabolic regulation (e.g., LEP, REN, MB), and immune modulation (e.g., CD38, CD99, MMP3). Consistent with this functional diversity, tissue-specific expression analysis based on consensus data from the Human Protein Atlas ^24^ and GTEx ^25^ revealed that 55 % of PSI proteins were enriched in specific tissue groups, rather than being broadly expressed across multiple tissues. This was significantly higher than the proportion observed among non-selected proteins (OR=5.37, *p*=4×10^−6^). The strongest enrichment was observed in male reproductive tissues, with 9 proteins showing specific expression in organs such as the testis and epididymis (**Figure 2c**).

**Fig. 2.**
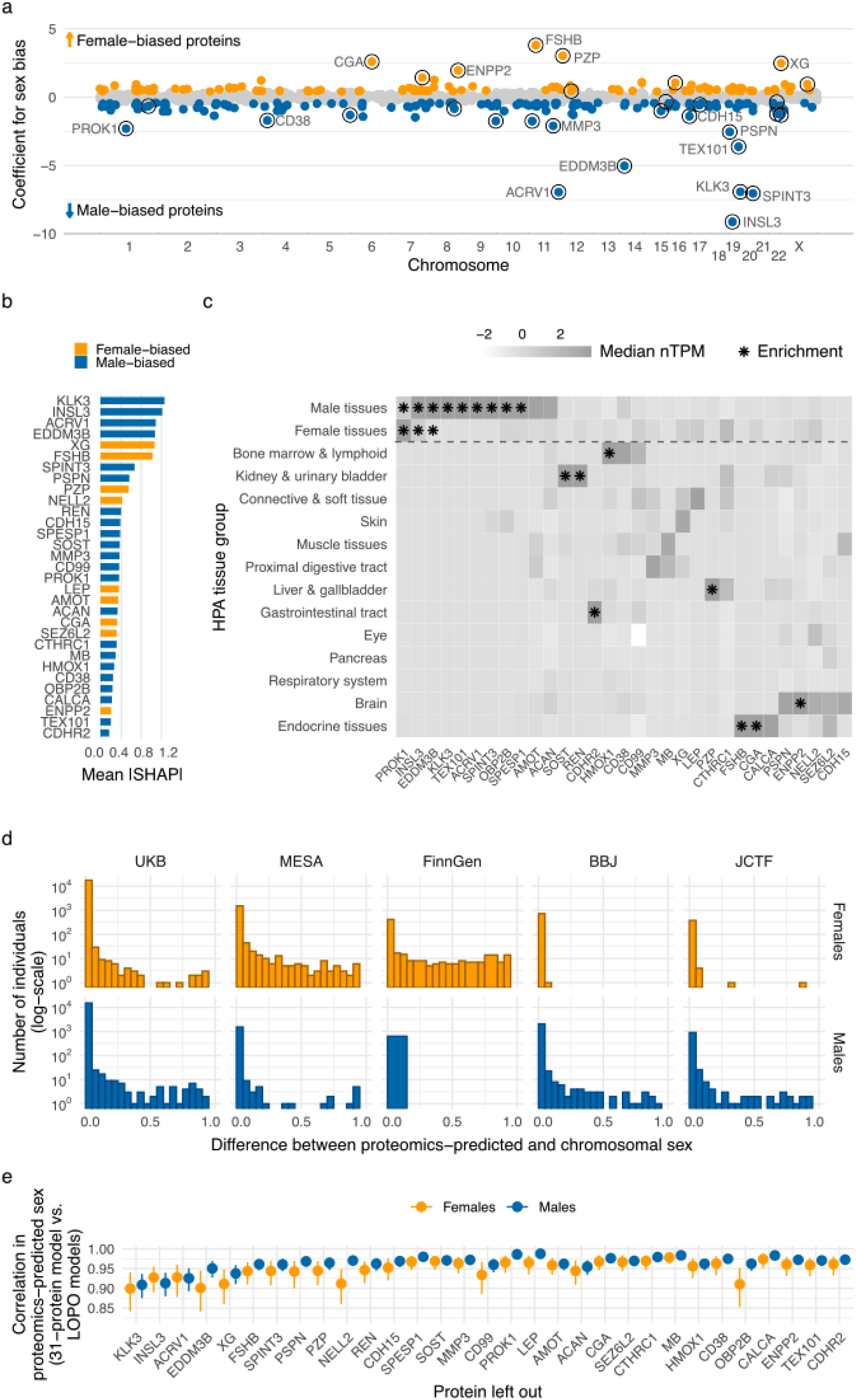
Proteomic sex index (PSI). **a**, Proteomic sex bias coefficients across the chromosomes based on the coding genes. Proteins with fold change >1.5 are highlighted (blue = male(XY)-biased, orange = female(XX)-biased). Sex bias is defined as the age-adjusted coefficient from a logistic regression between chromosomal sex and normalized protein expression (NPX) levels in the UK Biobank (N=42,979). Higher values in sex bias indicate female-biased proteins, i.e., proteins with higher expression in females, and lower values indicate male-biased proteins. Open circles indicate proteins included in the PSI model. **b**, Feature importances of the 31 proteins in the XGBoost model predicting chromosomal sex based on plasma protein expression measured using mean absolute SHAP values in the UK Biobank test set and annotated based on the direction of sex bias of each protein. **c**, Tissue group enrichment for genes encoding proteins included in the XGBoost model after feature selection (PSI proteins), normalized by gene. Stars (*) indicate tissue groups with at least 4-fold enrichment compared to any other tissue group, excluding female tissues when evaluating the enrichment in male tissues, and vice versa. **d**, Distributions of the distance between proteomics-predicted sex and chromosomal sex in the UK Biobank test set, MESA, FinnGen, BBJ, and JCTF, stratified by sex (orange = females, blue = males). **e**, Pearson correlations between predictions from the full 31-protein model and leave-one-protein-out (LOPO) models. Each protein is sequentially excluded, and a new XGBoost model is trained. The resulting predictions are correlated with those from the full model. The dots represent point estimates, and error bars denote 95 % confidence intervals. Leaving out any of the 31 proteins results in highly concordant predictions with the full model. Abbreviations: SHAP, SHapley Additive exPlanations; PSI, Proteomic Sex Index; nTPM, normalized transcripts per million; UKB, UK Biobank; MESA, Multi-Ethnic Study of Atherosclerosis; BBJ, Biobank Japan; JCTF, Japan COVID-19 Task Force; LOPO, leave-one-protein-out

While these 31 proteins combined could distinguish chromosomal sex very well (point-biserial correlation between chromosomal and proteomics-predicted sex r=0.9967, 95 % confidence interval (CI)=[0.9959, 0.9974]; see **Supplementary Table 5** for other performance metrics) in the UK Biobank test set (N=32,978), there were 125 males (XY sex chromosomes) and 79 females (XX sex chromosomes) for which the proteomics-predicted sex diverged significantly from chromosomal sex, with smaller changes observed for a greater proportion of individuals. Here, we define significantly divergent individuals as males with predicted male probability <0.95 and as females with predicted female probability <0.95. This suggests that while we can predict chromosomal sex from plasma protein expression with high accuracy, we do not do so perfectly, pointing towards remaining overlapping expression patterns between chromosomal sexes.

### Proteomics-predicted sex is not largely influenced by technical artifacts or individual proteins and replicates in four studies

We started by exploring whether technical artifacts could explain why some individuals clearly diverge from sex-typical proteomes (below the 95 % classification threshold), according to proteomics-predicted sex. To assess this, we examined the combined influence of factors related to sample processing and participant selection, including batch effects, assessment site, and overall protein expression levels in the UK Biobank. The combined effect of these potential technical artifacts explains 17 % of the variability in male (XY sex chromosomes) individuals and 11 % in female (XX sex chromosomes) individuals (**Methods**, **Supplementary Tables 6-8**), and could not be the sole explanation for deviations between proteomics-predicted sex and chromosomal sex. We computed proteomic principal components and confirmed that these discordant individuals were not separated by the first nine principal components (**Supplementary Fig. 2**). To further rule out sample swaps as a potential explanation, we applied Idéfix ^26^, a method that uses genetic information to detect mismatches between an individual’s genetically predicted and observed proteome, and confirmed that sample misassignment is unlikely to explain the observed variation in PSI (**Methods**, **Supplementary Fig. 3**).

Finally, we tested the robustness of proteomics-predicted sex to the selection of proteins in the model. First, we retrained the model by excluding all 61 proteins expressed in sex-specific tissues according to GTEx (**Supplementary Table 2**), 5 of which are included in the 31 proteins selected in the main model, to understand if we are mainly capturing biology specific to these tissues. We re-ran the SHAP-based feature selection exactly as it was described above, ending up with a model including 43 proteins (see **Methods**). The new model predicted chromosomal sex accurately (point-biserial correlation 0.996, 95% CI [0.995,0.997], **Supplementary Table 5**), and its within-sex predictions were correlated with the original model (Pearson r between model predictions within males r=0.529, 95% CI=[0.423,0.629]; within females r=0.421, 95% CI [0.280,0.543], **Supplementary Table 9**), suggesting that the main PSI model is capturing sex differences in protein expression extending beyond reproductive tissue-derived proteins. Second, we trained 31 leave-one-protein-out (LOPO) models and showed that no single protein was solely driving the model’s performance (**Figure 2e**, **Supplementary Table 9**). Third, we used previously published interplatform correlation results ^27^ to examine whether PSI proteins were robustly captured by both Olink and SomaScan ^28^ proteomic assays. Out of the 31 PSI proteins, 27 were measured by both platforms, and we saw stronger between-platform correlation for the PSI proteins compared to all proteins shared between the two platforms (median rank correlation ρ=0.48 *vs.* ρ=0.39). Out of proteins correlated between the two platforms (20 proteins; ρ>0.2), all showed a directionally concordant association with sex, with 11 of the associations being statistically significant (*p*<0.05) in SomaScan.

The model developed in UK Biobank showed strong performance in all four external studies (FinnGen: point-biserial correlation between proteomics-predicted and chromosomal sex r=0.946, 95 % CI [0.931, 0.959], MESA: r=0.962 [0.953, 0.971], BBJ: r=0.960 [0.950, 0.970], JCTF: r=0.938 [0.919, 0.956]; see **Supplementary Table 5** for other model performance metrics) supporting its robustness across different ancestry groups and study designs. Although the model performed well in predicting chromosomal sex in the external studies, it was not always well calibrated within each sex (**Figure 2d**). We show that this miscalibration is attributable to differences between studies in median protein levels within each sex. This likely reflects cohort-specific factors, including differences in the proportion of males and females, which affect normalization and the estimation of relative protein abundance (**Supplementary Fig. 4**; **Methods**).

To address this issue, we apply inverse rank-normalization to the proteomics-predicted sex to derive the PSI. This transformation also mitigates the impact of individuals with extreme values. Throughout the manuscript, we report results using PSI rather than the untransformed proteomics-predicted sex.

Taken together, these sensitivity analyses indicate that proteomics-based sex prediction generalizes well across external cohorts and ancestry groups, is not driven by any single protein, and that marked divergence towards proteomic profiles more common in the other chromosomal sex is unlikely to be due to technical artifacts.

### PSI captures key hormonally-driven life stage transitions and associates with sex-differential phenotypes within males and females

Given that age is a major determinant of protein abundance^11^, and that 12 of the 31 PSI proteins were previously included in a proteomic aging clock developed in the UK Biobank^29^, we evaluated the association between PSI and age. Although individuals were age-matched during model development, PSI remained associated with age in the UK Biobank and across replication cohorts (**Supplementary Table 10**). Nearly all PSI proteins showed significant associations (*p*<2×10^−3^) with age in both males (29/31 proteins) and females (27/31 proteins; **Supplementary Fig. 5**), consistent with a dynamic change of PSI across the lifespan.

To investigate age-related patterns earlier in life, we use proteomic data from 54 children aged 2-20 years and 1,276 adults aged 20-70 years, all measured using the Olink Explore HT panel in FinnGen. Because this panel lacks three PSI proteins (KLK3, CGA, MB), we trained a 28-protein PSI using the overlapping set (**Methods**), which showed good concordance with the original PSI in 38 adults profiled with both Olink Explore 3072 and Explore HT (r=0.42, *p*=0.008). Within children, PSI differed markedly from adult levels, with the largest differences observed prior to puberty (<12 years) and a shift toward adult-like values thereafter. Larger differences between pre-puberty and adult PSI were observed in children with XY sex chromosomes (OR=18.8, *p*=2×10^−4^, N=10) than in children with XX sex chromosomes (OR=3.15, *p*=4×10^−3^, N=13; **Supplementary Fig. 6)**.

In females, a strong age-related change was associated with menopause: 74 % of PSI-defining proteins were linked to menopause status in UK Biobank (**Supplementary Fig. 5**; **Supplementary Table 11**). Many of these proteins were also associated with oral contraceptive use before menopause and hormone-replacement therapy after menopause itself (**Supplementary Fig. 5**). PSI was significantly higher in females prior to menopause (age-adjusted β=-0.50, *p*=2×10^−68^; **Supplementary Table 10**; **Supplementary Fig. 6**), reflecting the age composition of the UK Biobank training set, in which 61.6 % of females had undergone menopause. To account for this, we trained two additional PSI models using females before and after menopause with age-matched males (**Supplementary Table 5**; **Methods**). The resulting PSI measures were highly correlated (r=0.842, *p*<2×10^−16^) but differed in protein contributions (**Supplementary Fig. 7**); for example, FSHB showed minimal sex differences before menopause but became strongly female-biased thereafter (**Supplementary Fig. 8**), and the differences in SHAP values between these two models reflect the same phenomenon. Both measures correlated with the primary PSI, with a stronger correlation observed for the postmenopausal-derived PSI (r=0.904) than for the premenopausal-derived PSI (r=0.758; **Supplementary Table 9**).

Next, we explored the association between PSI and three measures that differ on average between males and females: testosterone levels, height, and grip strength. The first two phenotypes were available in at least two studies, and we report the meta-analyzed results. Males with higher PSI, i.e., those with their proteome shifted towards females, had significantly lower testosterone levels, lower grip strength, and lower height. Conversely, females with higher PSI had higher testosterone, higher grip strength, and were taller (**Figure 3a**). These opposite directions indicate that protein levels more commonly observed in males are associated with higher values of these phenotypes in both sex groups. We observed, however, significant heterogeneity in height effect sizes for females (Cochran’s Q=15.18, *p*=0.004). In males, the association with height and grip strength did not significantly change after adjusting for total testosterone, and, in females, when adjusting for estradiol levels or menopause status. We additionally confirm that the association between PSI and testosterone was directionally concordant across total, free, and bioavailable testosterone, and that the association between PSI and height was largely unchanged after adjusting for these testosterone measures (**Supplementary Table 10**).

**Fig. 3.**
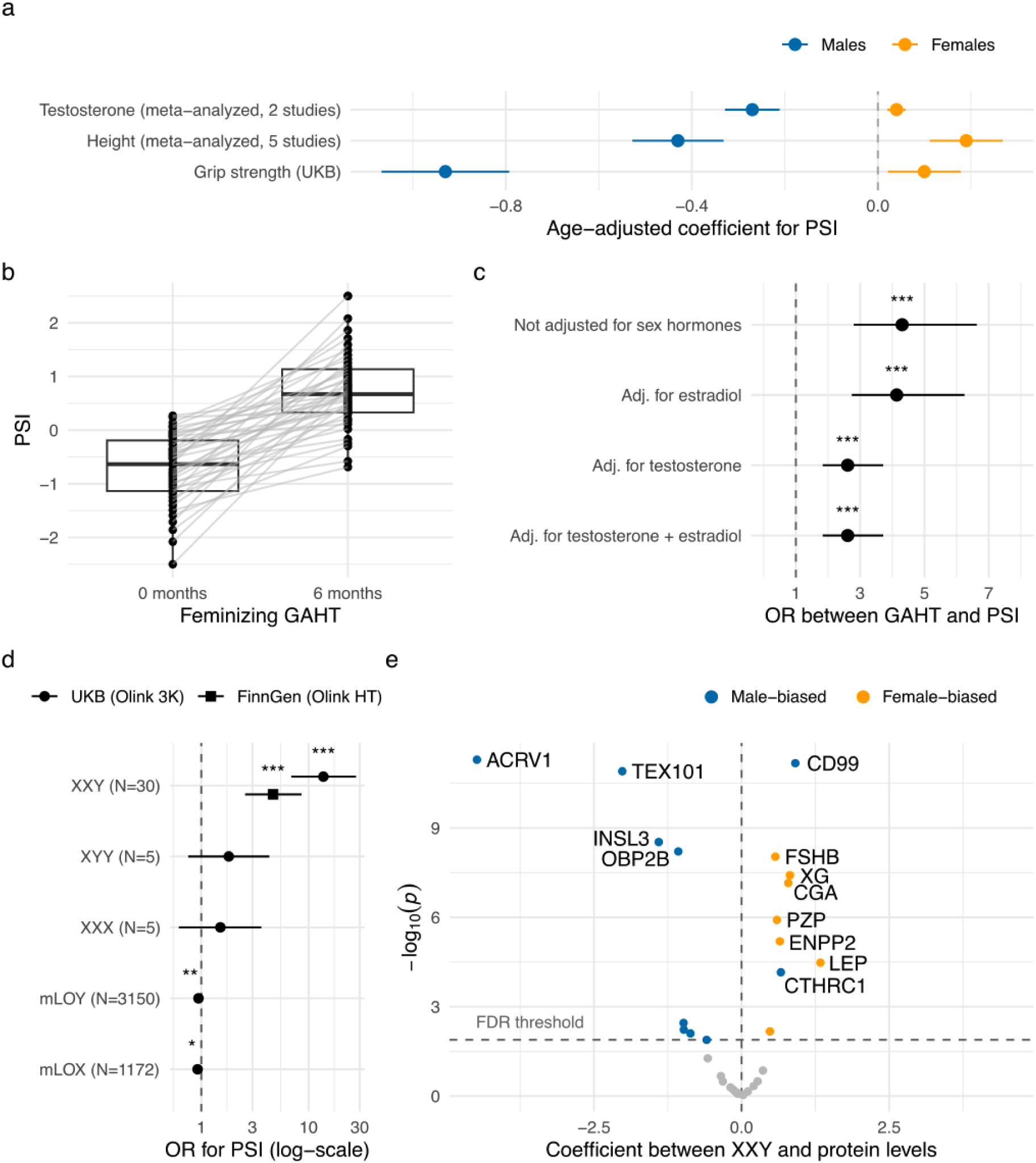
Association of PSI with sex-differential traits, gender-affirming hormone therapy (GAHT), and sex chromosome aneuploidies. **a**, Association between PSI and sex-differential traits. If results were available from multiple studies, we report the meta-analyzed associations. **b**, Boxplot of change in PSI before vs. after 6 months of feminizing GAHT. PSI is higher after GAHT, indicating a proteome shifted toward female-typical values. **c**, BMI- and age-adjusted association between PSI and GAHT. The association between PSI and GAHT persists when additionally adjusting for sex hormones. **d**, Odds ratio (OR) for the association of PSI with XXY (N=30), XYY (N=5), XXX (N=5), and somatic loss of X (N=1,172) and Y (N=3,150). Individuals with XX chromosomes were used as controls for XXX and mLOX, and individuals with XY chromosomes were used as controls for XXY, XYY, and mLOY. The association between PSI and XXY was replicated in FinnGen. PSI is strongly associated with XXY and slightly negatively associated with mLOX and mLOY. **e**, Volcano plot of the associations between XXY and each protein included in the PSI. The dashed line represents the FDR-significance threshold. Male-biased proteins (blue) tend to be negatively associated with XXY, while female-biased proteins (orange) tend to have a positive association. Abbreviations: UKB, UK Biobank; GAHT, gender-affirming hormone therapy; OR, odds ratio; mLOX, mosaic loss of X; mLOY, mosaic loss of Y; FDR, false discovery rate

We further confirm that single proteins within PSI were not driving these associations by repeating the analysis leaving out each protein at the time (**Supplementary Fig. 9**). All associations for height and testosterone remain statistically significant in the LOPO models in both chromosomal sexes, whereas for grip strength this holds for males and within females the effect size estimates are similar in magnitude but not always statistically significant.

Together, these findings indicate that PSI varies dynamically with age and captures key hormonally-driven life stages, including puberty in males and menopause in females. Nonetheless, after accounting for age and menopause status, PSI continues to capture meaningful within-sex variation in traits that show well-established differences between males and females.

### PSI changes during gender-affirming hormone therapy

We hypothesized that if PSI captures sex-related variation in the plasma proteome, it should change significantly in individuals undergoing gender-affirming hormonal therapy (GAHT). To test this, we utilized a previously published Australian cohort of N=40 transgender and non-binary individuals undergoing feminizing GAHT with antiandrogen therapy (cyproterone acetate or spironolactone) ^30^. Plasma protein expression was measured at baseline and after 6 months using the Olink Explore HT panel, and PSI was adapted to this platform using the same approach applied to the FinnGen pediatric cohort.

After 6 months, PSI increased in all individuals, indicating a shift in the proteome toward values commonly observed in XX individuals (meta-analyzed OR=4.30, *p*=3×10^−11^; **Figure 3b**), with similar results observed in both antiandrogen treatments (**Supplementary Table 12)**, independently of age and BMI changes. Notably, PSI changes remained significant after adjustment for circulating testosterone, estradiol, or both (**Figure 3c**), indicating that PSI captures GAHT-related biological changes not fully reflected by clinically measured hormone levels.

In the absence of data from a comparable longitudinal study of masculinizing GAHT, we generated proteomic data in FinnGen participants with indicators of GAHT initiation who had evidence of testosterone purchases and a biobank sample collected after the first purchase (**Methods**). In this group (N=23), PSI was statistically significantly elevated toward an XY-typical profile (OR=3.36, *p*=2×10^−5^; **Supplementary Table 12**).

### Sex-chromosomal trisomies and mosaic loss of X or Y associate with PSI

Sex chromosome trisomies, including Klinefelter syndrome (XXY), Triple X syndrome (XXX), and XYY syndrome (XYY), are among the most common genetic causes of differences in sex development. Using array intensity data from the UK Biobank, we identified 13 individuals with XXY and 5 individuals with XXX and XYY, each. We found a strong association between PSI and XXY so that one standard deviation increase in PSI among males (XY) was associated with a 13.7-fold increase (*p*=1×10^−13^) in the likelihood of having XXY (**Figure 3d**, **Supplementary Table 13**). We replicated this result in N = 17 XXY FinnGen participants (OR=4.68, *p*=6×10^−7^; **Supplementary Table 13**), with differences in effect sizes attributable in part to the use of a different Olink platform in FinnGen. Within both cohorts, we performed permutation tests and confirmed that the associations persisted (**Supplementary Table 13**). We further confirmed that the association with XXY persists in the LOPO models (**Supplementary Fig. 10**), suggesting this was not driven by a single protein. In fact, out of 31 proteins in PSI, 17 were significantly associated with XXY, spanning proteins both with and without sex-specific tissue enrichment, with female-biased proteins positively associated and male-biased proteins negatively associated (**Figure 3e**). The only major exception is for CD99 antigen (CD99), a male-biased protein that shows a strong positive association with XXY (OR=2.49, *p*=7×10^−12^). Located in pseudoautosomal region 1 (PAR1), *CD99* is partially silenced by X-inactivation in XX cells ^31^ and is overexpressed in XXY cells ^32^, likely explaining its opposite direction of association compared to the other PSI proteins.

We also considered mosaic loss of the X chromosome (mLOX) and the Y chromosome (mLOY) in the blood cells, an age-related phenotype primarily associated with cancer and cardiovascular diseases ^33,34^. PSI was negatively associated with mLOX (OR=0.93, *p*=0.01) and mLOY (OR=0.94, *p*=0.006) after adjusting for age and multiple testing correction (**Supplementary Table 13)**.

### PSI predicts the incidence of several sex-enriched diseases, including COVID-19, after adjusting for plausible risk factors

We explored associations between PSI and disease incidence in the UK Biobank using Cox proportional hazards models, adjusting for age, Townsend deprivation index, and covariates known to broadly influence protein expression: body mass index, serum creatinine, testosterone, and alanine aminotransferase levels (**Methods**). Among females (XX sex chromosomes), we additionally adjusted for menopause and hormone replacement therapy status. We considered 361 diseases defined by top-level Phecodes ^35^ with at least 20 incident cases and excluded individuals diagnosed with the disease before or within three months of the baseline proteomics sampling.

We observed more statistically significant associations (Bonferroni-corrected *p*<0.05) in females (19 diseases) than in males (6 diseases), with higher PSI consistently associated with increased disease risk (**Figure 4a**, **Supplementary Table 14**). Associations spanned multiple organ systems, and several, though not all, involved diseases known to differ by sex. For example, males with higher PSI—indicating a proteomic profile shifted toward values more typical for individuals with XX sex chromosomes—had an elevated risk of hypothyroidism (hazard ratio (HR)=1.25, 95% CI=[1.13, 1.39]), a disease approximately ten times more common in females than males in the UK ^36^. Similarly, higher PSI in males was associated with a greater risk of iron deficiency anemia (HR=1.26, 95% CI=[1.17, 1.37]), which is nearly twice as prevalent among females ^37^.

**Fig. 4.**
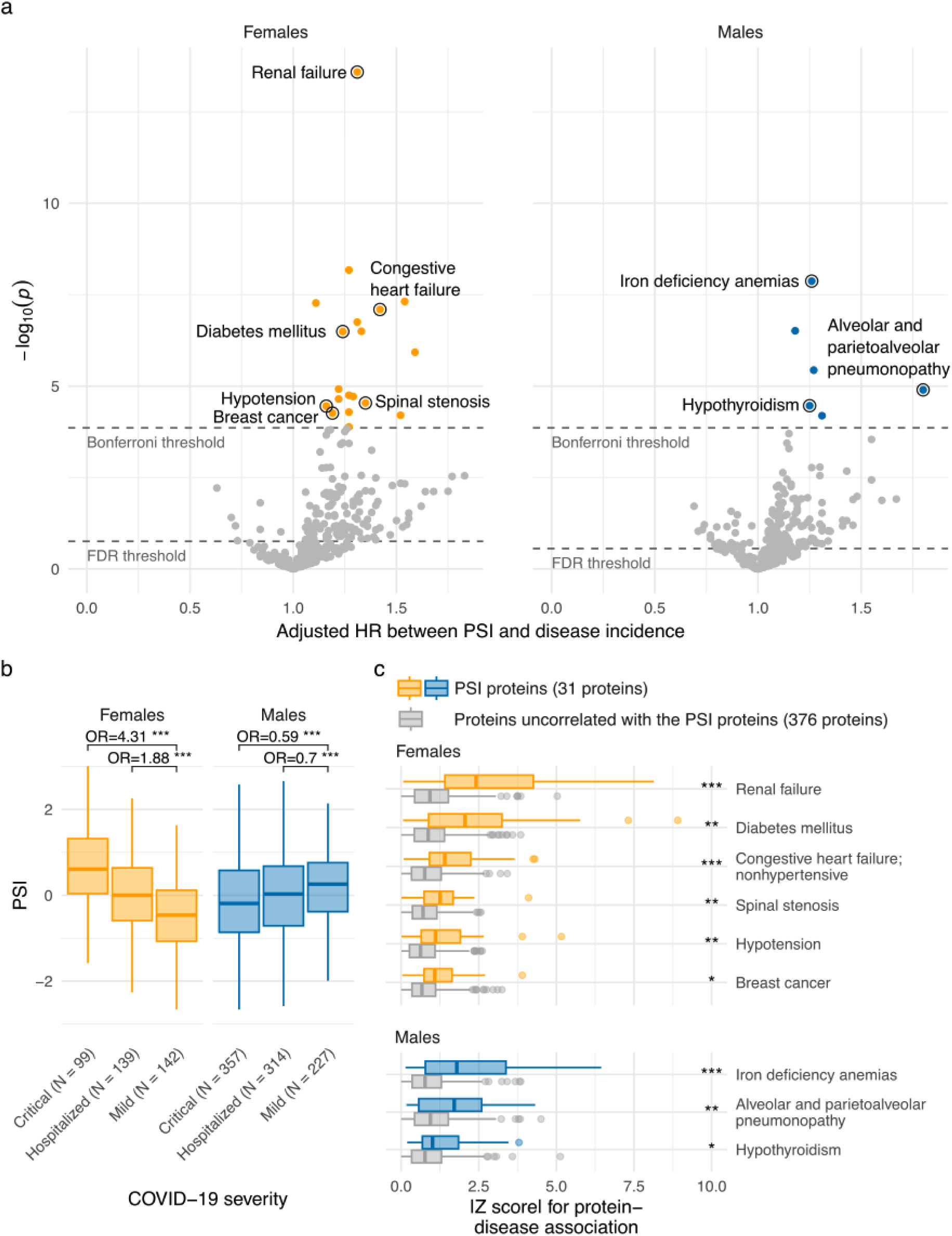
Phenotype and disease associations with PSI. **a**, Volcano plot of the association between PSI and disease incidence in the UK Biobank. Open circles denote the annotated diseases. **b**, Association between PSI and COVID-19 severity within JCTF (N=1,278). **c**, Significant protein-disease associations within selected PSI-associated diseases between PSI proteins (orange for females, blue for males; 31 proteins) and proteins not correlated (Pearson r<0.1) with the PSI proteins (gray; 376 proteins). Absolute z-scores represent scaled effect sizes for individual protein-disease associations, and stars represent statistical significance from Bonferroni-corrected one-sided Wilcoxon tests between the two sets of proteins (***, p<0.001; **, p<0.01, *, p<0.05). Abbreviations: UKB, UK Biobank; p, p-value; HR, hazard ratio; OR, odds ratio

Among females, the strongest associations with higher PSI were observed for renal failure (HR=1.31, 95% CI=[1.22, 1.41]), a disease more common in males ^38^. Higher PSI was also associated with increased risk of diabetes mellitus (HR=1.24, 95% CI=[1.14, 1.34]), a condition with slightly higher male prevalence in the UK ^39^. Other statistically significant associations, including nonhypertensive congestive heart failure, chronic skin ulcer, and hypotension, could plausibly reflect links to diabetes, but also to hormonal changes, given that estrogen is known to promote vasodilation and lower blood pressure. The significant association between higher PSI and breast cancer incidence (HR=1.19, 95% CI=[1.10, 1.30]) is notable, as breast cancer almost exclusively affects females.

Given the well-documented sex differences in COVID-19 severity, with males at higher risk of severe outcomes ^40,41^, we specifically examined associations between PSI and COVID-19 severity using data from the JCTF cohort ^20^. JCTF enrolled 1,372 individuals hospitalized with COVID-19–like symptoms in Japan (early 2020), of whom 493 were categorized as critical COVID-19, 484 as hospitalized, and 395 had mild disease. In this cohort, PSI was significantly associated with disease severity in both chromosomal sexes but in opposite directions (**Figure 4b**, **Supplementary Table 15**). In males, higher PSI, indicating a proteomic profile shifted toward values more typical of females, was associated with a reduced risk of critical COVID-19 (OR=0.59, *p*=3×10^−7^) after adjusting for age. In contrast, among females, higher PSI was associated with increased risk of critical disease (OR=4.31, *p*=1×10^−10^). Notably, PSI does not include proteins previously implicated in COVID-19 severity, such as ACE2 ^42^.

All analyses so far have been sex-stratified. However, most disease prediction models are applied to males and females and include sex as a predictor. While our goal is not to outperform disease-specific prediction models or to derive new prediction models based on PSI, we sought to assess whether PSI provides additional information for risk stratification beyond chromosomal sex. To test this, we compared a base model including binary sex and covariates used in the previous analysis with a model that additionally included PSI and its interaction with sex. The interaction term is necessary because PSI is constructed to have opposite directions of effect in the two sexes. The PSI augmented model showed improved fit for 71/290 diseases (ΔAIC > 4), with no diseases favoring the base model (ΔAIC < −4), significant likelihood ratio test improvement for 67 diseases (FDR-corrected *p*<0.05), and improved NRI risk reclassification for 29 diseases (**Supplementary Fig. 11**; see **Supplementary Table 16** for all metrics).

We performed multiple robustness analyses. First, we performed LOPO analyses for the 25 PSI-associated diseases and show consistent results, indicating that associations were not driven by single proteins (**Supplementary Fig. 12**). Second, because several diseases associated with PSI (e.g., renal failure) may induce proteome-wide changes, we tested whether the associations were specific to the 31 proteins included in PSI or could also be observed for unrelated proteins (**Methods**), allowing us to identify a subset of associations supported by enrichment among PSI proteins beyond proteome-wide disease signals. We observed significantly larger effect sizes for PSI proteins in 4/6 diseases in males and 8/19 diseases in females (one-sided Mann-Whitney U-test, Bonferroni-corrected *p*<0.05; **Figure 4c**; **Supplementary Fig. 13**).

These results show that high PSI is indicative of increased disease risk, largely for different diseases within males and females. Interestingly, many of these diseases have a higher prevalence in the other sex chromosome group.

### Identification of genetic factors associated with PSI and their impact on sex-enriched diseases and traits

Proteins included in the PSI can be elevated due to genetic effects (protein quantitative trait loci, pQTL), some of which may be sex-specific (sex-biased pQTL). These genetic effects may help explain part of the PSI variability observed within the chromosomal sexes. Supporting a genetic contribution to PSI, we observed a higher within-pair correlation of PSI values among monozygotic twin pairs (r_MZ_=0.75; 111 twin pairs) compared to same-sex dizygotic twins (r_DZ_=0.53; 79 twin pairs) in the Finnish Twin Cohort, yielding an estimated heritability of 43 % (**Methods**).

We conducted two genome-wide association studies (GWAS) of PSI in the UK Biobank, separately for males (XY sex chromosomes; N=14,077) and females (XX sex chromosomes; N=16,600), including only participants of genetically inferred European ancestry (**Figure 5a**). The autosomal SNP-based heritability of PSI was 0.183 (95 % CI [0.156, 0.210]) in males and 0.124 (95% CI [0.102, 0.146]) in females. We identified five independent loci associated with PSI: two genome-wide significant loci shared between the sexes (*MMP3* and *CDH15*), one female-specific (*PZP*), and two male-specific (*NELL2* and *XG*). All signals were cis-pQTLs for proteins included in the PSI model and exhibited opposite effects on PSI in males versus females (**Supplementary Table 17**). Overall, genes encoding PSI proteins were significantly enriched for sex-biased pQTLs (enrichment=13.9; *p*=5×10^−9^ using results from Koprulu et al. ^14^). To externally validate the GWAS findings—while accounting for the limited sample sizes of the available external cohorts—we constructed a composite score using one lead variant per locus. This score showed consistent direction and magnitude of association in both MESA and FinnGen (**Supplementary Table 18**).

**Fig. 5.**
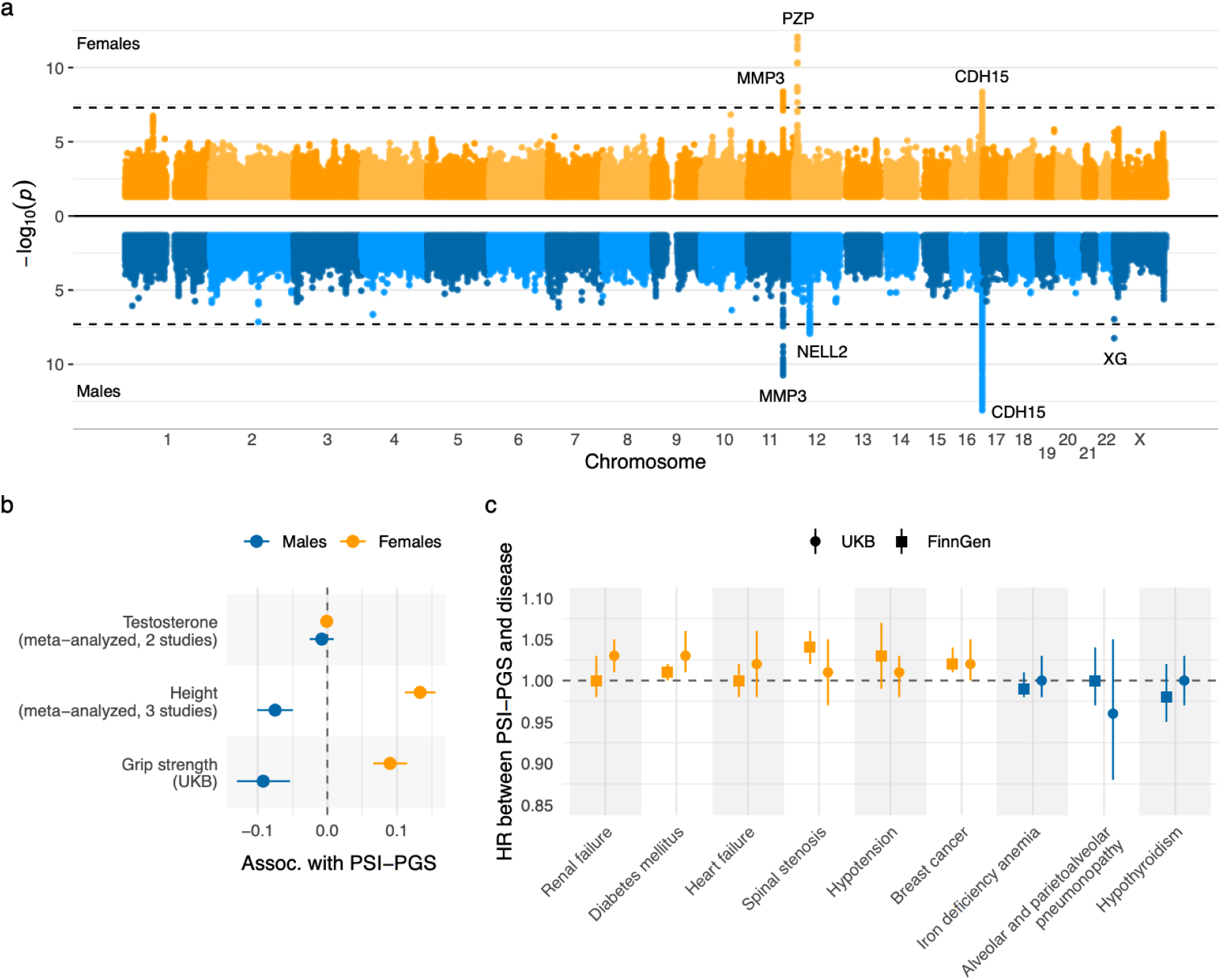
Genetic analysis of the Proteomic Sex Index (PSI). **a**, Miami plot showing genome-wide association results for PSI in females (top; N=16,600) and males (bottom; N=14,077) from the UK Biobank. The dashed line indicates the genome-wide significance threshold. Peaks are annotated according to protein quantitative trait loci (pQTL) found within the 31 PSI proteins. **b**, Association between PSI-PGS and sex-differential traits. If results were available from multiple studies, we report the meta-analyzed associations. **d**. Survival analysis to estimate the association between PSI-PGS and the first occurrence of diseases previously found to be phenotypically associated with PSI. Abbreviations: p, p-value; PGS, polygenic score; UKB, UK Biobank; HR, hazard ratio

Notably, the intronic variant rs2155013 at the MMP3 locus—associated with increased PSI in males and decreased PSI in females—was linked to a decreased risk of atherosclerosis in FinnGen (*p*=1×10^−8^; **Supplementary Table 17**). A missense variant in *PZP* (rs2277413) was associated with increased PSI exclusively in females, with the corresponding pQTL indicating reduced PZP levels. PZP is female-biased in the plasma, and reduced levels are consistent with a shift toward more male-typical protein levels. This variant was also linked to several female-specific clinical outcomes in FinnGen, including a lower risk of pregnancy hypertension (*p*=3×10^−5^) and failed induction of labor (*p*=1×10^−4^), underscoring its relevance to reproductive biology.

Using the PSI GWAS results, we constructed sex-specific polygenic scores (PGS) and tested their associations with sex-differential traits (i.e., height, testosterone, grip strength), as well as with diseases previously found to be phenotypically associated with PSI (**Figure 4b**; **Supplementary Table 10**). Where available, these analyses were replicated in external cohorts (FinnGen and MESA). The PSI-PGS was significantly associated with height and grip strength in directions consistent with phenotypic correlations. For example, males with a higher PSI-PGS were significantly shorter (**Figure 5c**; **Supplementary Table 20**). In contrast, no significant association was observed with testosterone levels, suggesting that the genetic determinants of PSI operate independently of testosterone regulation in adults.

In terms of disease outcomes, PSI-PGS was significantly associated with several conditions in females but not in males (**Figure 5d**; **Supplementary Table 21**). However, the effect sizes of the PSI-PGS associations with diseases were low. For instance, higher PSI-PGS in females was linked to increased risk of breast cancer (meta-analyzed HR=1.02, *p*=7×10^−5^) and type 2 diabetes (meta-analyzed HR=1.01, *p*=0.008).

Altogether, these analyses revealed that PSI has a heritable component, partially influenced by sex-specific genetic effect, and that genetically elevated PSI explains part of the associations observed with sex-enriched traits and diseases.

## Discussion

The main conceptual innovation of this study is the Proteomic Sex Index (PSI), a plasma proteomic indicator quantifying how closely an individual’s proteome aligns with their chromosomal sex. PSI captures continuous within-sex variation, allowing us to investigate sex differences independently of the sex chromosome complement. The development and application of PSI carry important implications. Sex is used as a key factor in medical decision-making, given that disease occurrence, progression, and treatment response often differ between males and females. PSI offers a nuanced and individualized measure of sex-related signatures, recognizing that some individuals may exhibit a proteomic signature more closely aligned with profiles commonly observed in individuals with different sex chromosomes.

We demonstrated that PSI is not an artifact of technical variability or driven by a few individual proteins, but rather reflects a wide range of biological processes, including reproductive biology, muscle development, cardiometabolic regulation, and immune modulation. If PSI reflects a continuum of chromosomal sex-related molecular variability, we hypothesized that: (1) it should correlate with sex-differential traits within each chromosomal sex; (2) gender-affirming hormone therapies should influence PSI; (3) a higher PSI in one sex might associate with diseases more common in the other sex chromosome group; and (4) genetic effects on PSI, including sex chromosome aneuploidies, should be partially sex-specific. Our extensive analyses supported these hypotheses, although not exclusively.

PSI was associated with traits that differ on average between males and females, including height, testosterone levels, and grip strength. Within both sex groups, individuals with higher PSI (indicative of a proteomic shift from the values typical to their sex chromosome group) had trait values more closely resembling those of the other sex, independently of circulating sex hormones, such as testosterone and estradiol, as well as menopausal status. PSI was also associated with diseases that differ in prevalence or severity between males and females. A notable example was COVID-19 severity: in the Japan COVID-19 Task Force cohort, higher PSI was associated with reduced severity in males but increased severity in females, suggesting that a protein signature more characteristically male-like is associated with increased severity in both sex groups. This pattern is consistent with the known higher severity of COVID-19 in males and suggests that PSI could enhance biological risk stratification beyond binary sex categories.

However, not all associations pointed exclusively to sex-related traits or diseases. PSI was also associated with diseases not traditionally considered sex-biased, suggesting that some proteins contributing to PSI may capture broader biological processes beyond conventional sex differences. One potential concern is that certain diseases have widespread effects on the proteome, and any proteomic index, regardless of its design, might capture such disease signals. Our comparison against unrelated proteins provided an additional specificity assessment of the disease associations. We also notice that for some sex-enriched diseases, the direction of PSI’s effect was not always as expected. For instance, females with a PSI shifted towards male-typical values were at increased risk of breast cancer, contrary to the anticipated lower risk. However, given that elevated testosterone has been shown to be a causal risk factor for breast cancer ^43^, the phenotypic association between PSI and higher testosterone within females might explain the direction of this association.

A central question remains: what exactly does PSI capture? Does it reflect intrinsic sex-specific biological processes, or does it also capture environmental or lifestyle factors that are stereotypically associated with sex and also influence the plasma proteome?

PSI likely reflects both. Because PSI proteins are selected based on sex differences in expression, any exposure that differentially affects protein abundance in males and females, including lifestyle, environmental, and socially mediated factors, may contribute to PSI variation.

At the same time, several lines of evidence support PSI capturing biologically meaningful sex-related variation. First, among transgender and non-binary individuals undergoing feminizing GAHT, PSI shifted substantially toward proteomic patterns more commonly observed in individuals with XX chromosomes after 6 months of hormonal therapy, independently of changes in circulating testosterone and estradiol, body mass index, and age. These findings highlight that proteomic profiles are dynamic and responsive to hormonal context, and suggest that PSI reflects aspects of this variation not fully captured by clinically measured hormone levels. Second, PSI was strongly associated with Klinefelter syndrome (XXY), with a one-standard-deviation increase in PSI corresponding to a 14-fold higher likelihood of an XXY karyotype compared with XY. This effect was replicated in FinnGen, albeit with a lower magnitude. Third, PSI was substantially heritable, indicating biological regulation of its components. Consistent with this, genetic associations with PSI implicated plausible causal links to disease risk; for example, in females, genetically predicted lower levels of PZP, corresponding to a shift toward male-typical PSI, were associated with lower risk of pregnancy complications. Finally, while PSI was associated with age, suggesting dynamic proteomic changes across the life course, these changes were not uniform. Marked shifts were observed in males before and after puberty, whereas in females, significant changes additionally occurred around menopause. These findings should be interpreted with caution, as our analyses were cross-sectional and PSI was derived in adults aged 40-70 years. While we could derive pre- and post-menopausal PSI scores and showed they were highly correlated, the limited number of children precluded analogous analyses.

Several limitations warrant consideration. First, although the Olink platform provides broad coverage of plasma proteins, it is targeted and may miss relevant proteins detectable through untargeted approaches such as mass spectrometry. Second, the proteomics-predicted sex probability distribution is very skewed; that is, most individuals have a proteome closely aligned with their chromosomal sex, and only few individuals have proteomes very similar to the other sex group. Inverse rank normalization, while enhancing robustness and avoiding issues with score calibration, inflates variation among typical individuals and compresses variation at the distribution tails. However, using a dichotomous threshold approach (i.e., classifying individuals above or below the 95 % probability threshold) yielded similar associations with height, grip strength, and testosterone (**Supplementary Table 10**). Third, PSI was trained to predict binary chromosomal sex, and our cohorts lacked information on gender identity, meaning we could not quantify the effect of gender expression and gendered behaviors in cisgender individuals. Thus, while PSI captures biological and environmental sex-related variation, it should not be interpreted solely as a marker of gender. Additionally, using a binary definition of sex in model training is a reductionist approach that overlooks both additional genetic variation in sex chromosomes and broader dimensions of sex beyond genetics. Fourth, while protein-level sex associations of PSI proteins are concordant across platforms, PSI has only been trained and evaluated on the Olink platform, and its performance on other proteomic technologies remains unknown. Finally, longitudinal studies are needed to determine how PSI shifts over time within individuals, and whether such changes relate to health outcomes. To reduce the risk of misinterpretation of the results and conclusions presented in this paper, we provide a general audience summary accompanying this paper (**Supplementary Materials**).

Our study demonstrates that the human plasma proteome exhibits a robust, continuous sex-aligned signature and shows its relevance for biological traits, disease risk, and genetic architecture. The conceptual framework underlying PSI could be extended to other molecular layers such as transcriptomics, metabolomics, and epigenomics, offering new avenues for characterizing chromosomal sex-related biological variation. For example, a recent study found specific cancer-related transcriptomic profiles associated differentially with sex-related gene expression, and suggested a continuous transcriptomics-based sex index could be used to personalize cancer treatment ^44^. In conclusion, PSI provides a quantitative measure of sex-related proteomic variation, providing new opportunities to study how within-sex molecular diversity contributes to human health and disease beyond binary sex classification.

## Methods

### Study participants

The UK Biobank is a large-scale biomedical database and research resource containing genetic, lifestyle, and health information from 502,505 individuals in the United Kingdom ^16^. Participants in the UK Biobank were aged 40 to 69 years at enrollment, with 54.1 % females. We restricted our sample to a subset of participants with antibody-based Olink Explore 3072 PEA proteomics data, measuring 2,923 unique assays ^15,45^. In the following, we refer to Olink Explore 3072 PEA data simply as (Olink) proteomics data, and highlight other proteomic platforms and panels separately. In the UK Biobank, we included individuals with proteomics data available for at least 2,500 proteins, after removing measurements not passing the UK Biobank quality control criteria, and individuals with missing sex information, leading to a final sample size of 42,978 individuals.

FinnGen ^17^ is a large-scale public-private research initiative that integrates genome and health registry data from over 500,000 individuals, primarily of Finnish genetic ancestry. Analyses were restricted to 1,199 samples measured by Olink Explore 3072 from the Blood Service Biobank, representing a relatively healthy segment of the population. In our genetic analyses, where proteomics data were not required, we utilized the full FinnGen R12 cohort. We further included Olink Explore HT samples from blood donors (N=1,413, 38 with proteomics data measured from the same blood draw in both Olink Explore 3072 and Olink Explore HT), and generated new proteomic data from adolescents (N=54), individuals with Klinefelter syndrome (XXY; N=17), and individuals with evidence of masculinizing GAHT (N=23).

The Multi-Ethnic Study of Atherosclerosis ^18^ (MESA) is a longitudinal study designed to investigate cardiovascular disease and its progression and includes data from 6,814 individuals free of cardiovascular diseases at baseline. Our primary analyses focused on the MESA Exam 1 (data collected 2000–2002). After excluding samples not passing the quality control criteria and individuals with discordant self-reported and genetic sex, the baseline cohort comprised 2,851 participants.

The Biobank Japan ^19^ (BBJ) project is a hospital-based cohort study that recruited approximately 200,000 patients with complex diseases across multiple hospitals in Japan, representing a population enriched for diseased individuals. We included the subset of individuals with proteomics data available and concordant genetic and self-reported sex, comprising 2,870 individuals.

The Japan COVID-19 Task Force ^20^ (JCTF) was established in early 2020 as a nationwide multicentre consortium to overcome the COVID-19 pandemic. We included a subset of 1,372 individuals with proteomics data available and concordant genetic and self-reported sex.

Additionally, we assessed the effect of antiandrogen-specific remodeling of the plasma proteome to PSI in a previously published Australian cohort of 40 transgender and non-binary individuals undergoing feminizing gender-affirming hormonal therapy (GAHT) ^30^. In this cohort, plasma protein levels were measured for 5,279 proteins using the Olink Explore HT panel at baseline before the start of the antiandrogen therapy, and at 6 months following the start of the therapy. Two groups of GAHT were included: one with estradiol plus antiandrogen CPA, and the other with estradiol plus antiandrogen SPIRO, with both containing 20 participants.

### Protein measurements and preprocessing

The protein expression data in each of the main cohorts – UK Biobank, FinnGen, MESA, Biobank Japan, and Japan COVID-19 Task Force – were obtained using the Olink Explore 3072 PEA assay, employing the Proximity Extension Assay (PEA) technology. Protein expression levels were reported as Normalized Protein Expression (NPX) units on a logarithmic scale. We considered all proteins available in the UK Biobank, with the exception of four proteins (GLIPR1, NPM1, PCOLCE, and CTSS) being excluded due to missing measurements in over 20% of the UK Biobank sample. This results in a final set of 2,919 proteins.

Across the replication cohorts, we excluded any proteins not within this set of proteins and used the MissForest package ^46^ in Python to impute missing values in the other proteomic measurements. Within the FinnGen cohort, four proteins (CTAG1A_CTAG1B, EBI3_IL27, TNFSF9, and TOMIL2) were additionally excluded due to complete missingness among the selected participants, and 16 proteins unique to batch 1 were also removed. We trained the imputation models separately in each cohort and imputed a single dataset per cohort using a maximum of five iterations. All other parameters were left at default values. In the UK Biobank, the imputation model was trained on 10,000 individuals randomly selected from the full dataset. In the replication cohorts, the imputation models were trained using all available individuals.

Additionally, to account for technical variation across batches in the MESA proteomics data, we batch-corrected the proteomics data by applying linear regression to each protein. We modeled the NPX values as a function of age, sex, ancestry group, and batch indicator, and obtained the batch-corrected NPX values by subtracting the estimated batch effect from the NPX.

In addition to the Olink Explore 3072 PEA data, we utilized two cohorts where protein expression was measured using Olink Explore HT. In the Australian GAHT cohort, 5,279 proteins were measured, with exactly the same preprocessing as described previously ^30^. No imputation was required. Within FinnGen, we generated new proteomics data using Olink Explore HT as follows. Approximately 7 ml of venous blood was collected for research in EDTA tubes. For all samples, the plasma component was separated from cellular components by centrifugation (10 min at 500g, room temperature). Plasma samples were aliquoted and stored at −80 °C until analysis. Plasma samples were thawed, plated, and shipped to GeneWiz (Azenta Life Sciences) to assess 5,328 proteomic biomarkers with the antibody-based Olink Explore HT. The Explore HT platform utilizes Olink’s PEA technology, where multiplexed oligonucleotide-labeled antibody probe pairs attach to their target protein in the sample. Protein expression was quantified by next-generation sequencing. Protein concentrations were expressed as the relative concentration unit normalized protein expression. Values were log2-transformed to account for heteroskedasticity. Samples failing standard quality control criteria were excluded, with the exception of samples flagged as sex outliers by the FinnGen pipeline, which were retained if they passed all other QC metrics. These samples comprised children, individuals with sex chromosome trisomies, and samples with indicators of gender-affirming hormone therapy, and the sex outlier flags are defined relative to protein expression patterns typical of adults with no sex chromosome aneuploidies or hormone treatment, and therefore do not necessarily indicate technical failure in these groups. NPX values were normalized by fitting a scaling transformation in FinnGen blood donors previously provided with Olink Explore HT and applying the same scaler to the newly generated samples.

### Definition of sex

In the UK Biobank, sex was determined using chromosomal sex (UKB Data-Field 22001; ‘Genetic sex’), which provides sex based on genotyping analysis. In all replication cohorts, chromosomal sex was inferred using PLINK ^47^ based on X-chromosome heterozygosity (F-statistic). Females (XX sex chromosomes) are expected to show high heterozygosity (F-statistic close to 0), whereas males (XY sex chromosomes) are expected to appear largely homozygous (F-statistic close to 1).

We identified the sex chromosome trisomy status in the UK Biobank similarly to how it was previously reported ^48^. Shortly, we analyzed raw signal intensities from single-nucleotide polymorphism (SNP) array data on chromosome X and chromosome Y separately. For every UK Biobank participant, we first calculated log-R ratios (LRR) for each SNP located on the sex chromosomes and then obtained the individual-level chromosome X LRR and the chromosome Y LRR by taking the median value across all examined chromosome X SNPs and chromosome Y SNPs (i.e., median LRR). In calculation of LRRs, we corrected for the signal intensities of the autosomes in order to systematically exclude false positive trisomy detection resulting from sample contamination. We considered individuals with detectable Y chromosome intensity signals to be chromosomal males and those without to be chromosomal females. Clustering thresholds into five major sex chromosome karyotypes – 46,XY, 47,XXY, and 47,XYY for genetic males and 46,XX and 46,XXX for genetic females – have been previously described ^48^.

### Identification of sex-biased protein expression

We used a logistic regression model to evaluate the association between each protein and sex, which we call the sex bias of each protein. Sex bias is an age-adjusted regression coefficient between each protein and chromosomal sex, with positive values indicating higher protein expression in females (XX sex chromosomes) compared to males (XY sex chromosomes; female-biased proteins) and negative values indicating higher expression in males (male-biased proteins). Proteomics data were z-standardized before fitting the model, and the models were adjusted by age at recruitment. We used Bonferroni correction to account for multiple testing (Bonferroni-corrected p-value threshold 2×10^−5^). We assessed the concordance in sex-bias estimates across cohorts by computing Pearson and Spearman correlations between the sex bias estimates from UK Biobank and each of the replication datasets.

### Tissue enrichment of protein-coding genes

We investigated the tissue enrichment of the genes encoding proteins included in the analysis using tissue-specific median gene expression data from the Genotype-Tissue Expression ^25^ (GTEx; release 10, version 2.4.2) and Human Protein Atlas ^24^ (HPA; version 24.0) databases. This approach allowed us to infer tissue-specific expression based on gene expression profiles in various tissues, despite proteins being measured only in plasma. We used the HPA and GTEx consensus transcript expression levels in 50 tissues, mapped into 15 tissue groups according to the grouping used by the HPA. Importantly, epididymis, prostate, seminal vesicle, and testis were grouped as “male tissues” and breast, cervix, endometrium, fallopian tube, ovary, placenta, and vagina were categorized as “female tissues”, consistent with the HPA definitions. We excluded tissue groups with fewer than 10 enriched genes. We considered genes as tissue-enriched if the maximum median transcripts per million (TPM) of the tissue group was at least 4x that of any other tissue group. We also conducted the tissue enrichment analysis using GTEx data only, and the results of both analyses are provided in the **Supplementary Table 2**.

### Training the proteomics-based prediction model for chromosomal sex

We used UK Biobank proteomics data to develop a proteomics-based predictor of chromosomal sex. We first divided the UK Biobank proteomics dataset into training (20 %) and test (80 %) subsets after performing imputation.

We applied additional filtering to the training set only. We began by excluding individuals with discordant chromosomal and self-reported sex, as well as those with sex chromosome aneuploidies (i.e., individuals with sex chromosomes other than XX or XY; UKB Data-Field 22019; ‘Sex chromosome aneuploidy’). To minimize the risk of the model learning age differences between sexes in the UK Biobank, we then matched the age distributions of males and females. This was done by subsampling the data such that each one-year age bin (UKB Data-Field 21022; ‘Age at recruitment’) contained an equal number of males (XY sex chromosomes) and females (XX sex chromosomes). Individuals who were not included in the age-matched subset were excluded from both the training and test sets. Following this, we normalized (z-standardized) the NPX values by estimating the scaling factors from the training set and then applying the same transformation to both the training and test sets.

After these preprocessing steps, the final training set consisted of 9,128 individuals. The test set, which remained unaltered aside from the initial imputation and normalization, comprised 32,978 individuals and was used to evaluate model performance and conduct all downstream analyses.

We next performed feature selection by further dividing the training set into 85 % used for updating the model parameters and 15 % used for validation and hyperparameter tuning. We trained 1,000 extreme gradient boosted decision tree models using the XGBoost library ^21^ (version 1.6.2) to predict chromosomal sex (XX or XY) based on all 2,919 proteins as features, inspired by the Stochastic Gradient Boosting (SGB) ensembles approach ^23^. In this approach, we optimize the XGBoost hyperparameters ‘subsample’ and ‘colsample_bytree’ (search space: 0.5 to 0.95; 20 equally spaced values) during training, initializing each model with a different random seed, which introduces variation into the learned model parameters.

During training, we performed Bayesian hyperparameter optimization using the Skopt library ^49^ (version 0.10.1). The search space included: ‘learning_rate’ (0.0001, 0.001, 0.01, 0.1, 0.2, 0.3), ‘n_estimators’ (100, 300, 800), ‘max_depth’ (3, 5, 6, 10), ‘gamma’ (0 to 15; 20 equally spaced values), ‘reg_lambda’ (1 to 20; 10 equally spaced values), and ‘reg_alpha’ (fixed at 0). All other hyperparameters were left at their default setting.

Feature importances were estimated using absolute SHapley Additive exPlanations ^22^ (SHAP) values (SHAP package version 0.42.1). From the ensemble of 1,000 models, we selected protein features whose 95 % confidence intervals for |SHAP| values exceeded 5 % of the mean |SHAP| value of the most important protein (*KLK3*). The 95 % confidence intervals were computed over 1,000 bootstrap samples drawn from among the |SHAP| values of each of the models in the ensemble for each protein. This approach resulted in 31 protein features, which are listed in **Supplementary Table 4**.

We then trained a final model using only these 31 proteins as input features. Hyperparameter optimization was performed as described above, with the exception of fixing ‘subsample’ and ‘colsample_bytree’ to 1 to eliminate stochasticity in the final model. A schematic overview of the training pipeline is provided in **Figure 1**. The XGBoost models are binary classifiers and output the predicted male probability (XY sex chromosomes) P_XY_ that varies between 0 and 1.

To compute PSI in the GAHT cohort and within the FinnGen samples with proteomics measured using the Olink Explore HT panel, we re-trained a 28-protein PSI with the three main PSI-proteins (KLK3, CGA, MB) that are not measured in the Olink Explore HT panel removed. After this, the training was done similarly to the main PSI. We then confirmed that this 28-protein PSI is highly correlated (Pearson correlation) with the main PSI both in the UK Biobank test set with proteomics measured using Olink Explore 3072 and in a subset of FinnGen participants for which we had both Olink Explore 3072 and Olink Explore HT measurements from the same blood draw available.

A sensitivity analysis using the same training pipeline was conducted by allowing selection of the model features from among those proteins whose coding genes were not significantly enriched in sex-specific tissues based on GTEx. This restriction removes many of the top features from the 31 protein model, such as KLK3 (rank 1 according to |SHAP|), INSL3 (rank 2), ACRV1 (rank 3), and EDDM3B (rank 4). After the feature selection pipeline, this model includes 43 proteins: XG, FSHB, PZP, PSPN, LEP, CD99, CGA, ENPP2, MMP3, CD38, SEZ6L2, CDHR2, OBP2B, NPTX2, MB, CDH15, ACE2, AMOT, WFDC12, CTHRC1, SERPINA6, RECK, SPRR3, IGFBP6, SCRG1, TSPAN1, VIT, ENDOU, CALCA, PAEP, SHBG, APCS, IGFBP3, ACAN, NCAN, REN, ADGRD1, POMC, ADGRG2, SOST, THBS2, IL3RA, and FCGR3B.

To assess the effect of menopause on PSI, we additionally trained pre-menopause and post-menopause PSI models in UKB. We used the same 31 proteins included in the main PSI, but subsampled the training populations so that they include only females before menopause and females after menopause, respectively, as well as age-matched males. The training set males were age-matched within each group separately. After this, the pre-menopause PSI and the post-menopause PSI were trained similarly to the main PSI.

To assess the stability of the final 31 protein model, we also trained 31 additional models, where each of these models was only allowed to use 30 proteins as features, leaving out one protein at a time. We call these leave-one-protein-out (LOPO) models.

### Model evaluation

Model performance was assessed on the independent UK Biobank test set (N=32,978) and across all replication cohorts using four complementary metrics: (1) the point-biserial correlation between predicted probabilities and genetic sex (encoded 0 for females and 1 for males) to quantify linear association; (2) the area under the receiver-operating-characteristic curve (AUC); (3) the area under the precision–recall curve (AUPRC); and (4) the Brier score. Ninety-five percent confidence intervals for all metrics were calculated via 2,000 bootstrap resamples within each cohort. These same metrics were also computed for each LOPO model, the 43-protein model excluding proteins coded by genes enriched in sex-specific tissues, the 28-protein model excluding proteins not included in the Olink Explore HT panel, and the two menopause status-specific PSI models.

### Proteomic sex index (PSI)

To quantify the extent to which an individual’s protein levels resemble those of the other chromosomal sex, we extracted the predicted male probability P_XY_ from the trained XGBoost model and transformed it into predicted distance from the chromosomal sex

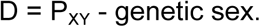

Where genetic sex is coded as 0 for females (XX sex chromosomes) and as 1 for males (XY sex chromosomes). These distances were then inverse rank-normalized within each chromosomal sex to remove distributional differences between sexes. The resulting metric, which we term the Proteomic Sex Index (PSI), captures an individual’s relative proteomic divergence from the profile typical of individuals sharing the sex chromosome group with them. Higher PSI values indicate greater deviation from the expected proteomic pattern for that individual’s chromosomal sex.

### Evaluating potential technical artifacts and proteomic contributions to extreme PSI values

To assess whether individuals with extreme PSI values were enriched for potential technical artifacts, we performed logistic regression comparing the top 1 % of PSI values to the remaining 99 % within the test set, separately for each sex. The selected quality control variables included assessment center (UKB Data-Field 54; ‘UK Biobank assessment centre’; categorical), UK Biobank Pharma Proteomics Project selected participant status (UKB Data-Field 30903; ‘UKB-PPP selected participant’; binary), date of attending the assessment center (UKB Data-Field 53; ‘Date of attending assessment centre’, modeled as days since the first visit), sample plate (UKB Data-Field 30901; ‘Plate used for sample run’; categorical), sample well (UKB Data-Field 30902; ‘Well used for sample run’; categorical), and mean NPX value across all 2,919 proteins. Categorical variable levels with fewer than 3 individuals with extreme PSI values were omitted. The retained levels in each analysis are presented in **Supplementary Table 6**. We then quantified the contribution of these technical variables to extreme PSI values by calculating the proportion of variance explained (pseudo-R^2^) from logistic regression models, separately for the 31-protein and 43-protein PSI definitions and within each chromosomal sex.

We further assessed whether individuals with extreme PSI values could be distinguished based on proteomic principal components. We performed principal component analysis on the NPX matrix of all 2,919 proteins after standardizing each protein to zero mean and unit variance, separately for XX and XY sex chromosome groups. We then assessed whether individuals in the top 1 % of PSI values could be distinguished from the rest of the population based on the first 9 principal components.

### Detecting potential sample swaps with Idéfix

We applied Idéfix ^26^, a method that uses genetic information to detect mismatches between an individual’s genetically predicted and observed phenotypes, to identify potential sample swaps within the UKB proteomics dataset. We first computed proteomic polygenic scores (pPGS) for UKB participants of European ancestry (N=39,880) using proteomic genome-wide association study (GWAS) summary statistics from the INTERVAL ^50^ cohort, available via the OmicsPred ^51^ database. We considered proteins measured by the Olink platform in the UKB (2,919 proteins) and obtained corresponding GWAS summary statistics from the OmicsPred database, using results derived from either Olink (302 proteins) or SomaScan ^28^ (989 proteins) platforms in the INTERVAL cohort, given the limited availability of Olink-based summary statistics within the database. We used PLINK2 to derive the pPGS. We then estimated the proportion of variance in observed protein levels explained by the pPGS and filtered proteins to retain those with an R^2^>0.2, using results from a platform with the higher R^2^ in cases where the same assay is included in both platforms. We further excluded highly correlated proteins (absolute Pearson correlation>0.5), resulting in a final set of 94 proteins. Notably, these proteins do not overlap with the 31 proteins used to train PSI. We then ran Idéfix using the observed protein levels and corresponding pPGS for each individual. The resulting scaled log-likelihood ratios represent how aligned an individual’s proteome is with their genetics, with higher values indicating greater deviance and a higher likelihood of a sample mismatch. Finally, we compared these scores to PSI values to assess whether individuals with extreme PSI values are more likely to deviate from their genetically predicted proteome.

### Evaluating the cross-platform sex bias

To explore the robustness of the PSI proteins across different proteomic platforms, we used previously published results ^27^ comparing protein measurements from Olink Explore 3072 and SomaScan 11K. We used UniProt IDs to link PSI proteins to proteins reported in the cross-platform dataset, and restricted the analysis to proteins shared between the two platforms. For proteins represented by multiple analytes in SomaScan 11K, we calculated protein-level medians. We compared cross-platform correlations of PSI proteins with all proteins shared across the two platforms by computing the median correlation. Within PSI proteins with high cross-platform correlation (Spearman ρ > 0.2), we extracted the reported associations with sex and compared their directional concordance across the two platforms.

### Quantifying sex-differential changes in median protein levels across cohorts

To investigate whether cohort-level differences in protein expression could contribute to sex-differential model calibration, we analyzed median NPX values for the 31 proteins used in the PSI model. For each protein, we computed the chromosomal sex-specific median NPX in each replication cohort (BBJ, JCTF, MESA, and FinnGen) and compared these to the corresponding medians in the UK Biobank training set. This analysis was motivated by the hypothesis that, despite normalization, cohort-level changes in median NPX levels towards those typical of the other sex could potentially result in the model being more likely to falsely classify cohort-wise typical values, i.e., individuals at median NPX for that cohort, as shifted towards the other sex. Conversely, shifts in the median values away from the other sex could result in the model being more likely to classify those as sex-typical values. To assess this, we calculated the absolute difference in median NPX between each cohort and UK Biobank, separately for males (XY sex chromosomes) and females (XX sex chromosomes), and determined whether the larger shift in each protein was directed toward or away from the other sex median in UK Biobank.

### The effect of age, puberty, and menopause

We estimated the associations between PSI and age in the UK Biobank, FinnGen, MESA, Biobank Japan, and JCTF using unadjusted linear regression models, using PSI as the outcome. Protein-age associations were tested separately in males and females using linear regression with PSI or the protein levels as the covariate and age as the outcome in the UK Biobank, applying Bonferroni correction to account for multiple testing.

In the UK Biobank, we computed the associations between PSI and PSI-defining proteins with menopause status (UKB Data-Field 2724; ‘Had menopause’; categories ‘Yes’ and ‘No’ included), current oral contraceptive use (UKB Data-Field 2804; ‘Age when last used oral contraceptive pill’; modeled as binary), and hormone-replacement therapy (UKB Data-Field 2814; ‘Ever used hormone-replacement therapy (HRT)’; categories ‘Yes’ and ‘No’ included) within females. We included only females before menopause in the oral contraceptive analysis and females after menopause in the HRT analysis. Associations were tested using linear regression, adjusting for age, and we applied Bonferroni-correction to account for multiple testing.

To assess the effects of menopause on sex differences in protein levels, we computed sex bias estimates within females aged 45 to 55 at the time of recruitment and age-matched males in the UK Biobank. Within this age range, we first matched the age distributions between males and females to control for age-related confounding, and further downsampled the larger group to match the sample sizes between the ‘Yes’ and ‘No’ menopause groups. Sex differences in protein levels were then assessed within each matched subset by computing the z-scores representing the scaled differences in sex bias magnitude. To evaluate the overall shifts in sex bias before and after menopause, we also calculated the mean absolute sex difference across all proteins and within the set of PSI-defining proteins, comparing these values between the menopause groups. We applied Bonferroni-correction to account for multiple testing.

To characterize PSI patterns earlier in life and over the menopausal transition, we analyzed plasma proteomic data from 54 FinnGen individuals aged 2-20 years and 1,276 adults aged 20-70 years, measured using the Olink Explore HT platform. We used proteomic data from all FinnGen blood donors with proteomics from Olink Explore HT to normalize the protein expression to mean 0 and standard deviation 1, and computed PSI using the 28-protein PSI model within these individuals. Because FinnGen does not include direct information on menopausal status, we used FSHB protein levels, which we saw rises markedly after menopause in the UK Biobank, to classify females as being before or after menopause. Because the FSHB distribution in females exhibited the same bimodal distribution within females as in the UK Biobank, with the lower peak aligning with that of males, we assumed that this protein could be used as a proxy for menopausal transition. We estimated the FSHB NPX cutoff between the two FSHB-estimated menopausal groups by fitting a 2-component Gaussian mixture model to females’ data, and classified females with FSHB levels below this threshold as being before menopause and those above the threshold as being after menopause. We then compared the PSI distributions across age bins using linear regression models comparing PSI between the groups and with two-sided Wilcoxon rank-sum tests.

### Associations of PSI with sex-differential phenotypes

We examined the association between PSI and height, testosterone, and grip strength. Analyses were performed separately within chromosomal sex groups XX and XY, and adjusted for age. In the UK Biobank, BBJ, and JCTF, we additionally excluded individuals whose predicted sex was fully discordant with chromosomal sex (classification threshold = 0.95), to minimize the impact of potential sample misannotation or technical artifacts. Outcomes were analyzed in their original (non-standardized) units using linear regression within each cohort. For height and testosterone, which were available in multiple cohorts, we performed fixed-effects meta-analysis. Heterogeneity across cohorts was assessed using Cochran’s Q statistic. As a sensitivity analysis, we repeated the models with additional adjustment for testosterone in both chromosomal sex groups and for oestradiol and menopausal status in females. We also performed leave-one-protein-out analyses, re-computing PSI with each of the 31 model proteins excluded in turn, to assess whether associations were driven by any single protein.

In addition to the total testosterone concentrations directly provided by UK Biobank and MESA, we also considered free and bioavailable testosterone, which are considered more clinically relevant under the free hormone hypothesis ^52^. In circulation, most testosterone is tightly bound to sex hormone-binding globulin (SHBG), while a smaller fraction is weakly bound to albumin and a minor fraction remains unbound (“free”). The albumin-bound and free fractions are collectively referred to as bioavailable testosterone. In MESA, both free and bioavailable testosterone were directly measured. In UK Biobank, these were estimated from total testosterone (UKB Data-Field 30850; ‘testosterone’), sex hormone-binding globulin (UKB Data-Field 30830; ‘SHBG’), and albumin (UKB Data-Field 30600; ‘albumin’) concentrations using the Vermeulen equation ^53^.

### PSI change in individuals undergoing feminizing gender-affirming hormonal therapy (GAHT)

Protein abundances in the GAHT were measured using the Olink Explore HT panel. Thus, we used the 28-protein PSI model, which includes only those 28 main PSI proteins that are included in the Olink Explore HT panel. PSI values were computed in the GAHT cohort as follows. The NPX values were normalized (z-standardized), with both time points normalized together. No imputation was required. After this, PSI was computed similarly to the other cohorts.

To assess the change in PSI at 6 months after initiation of GAHT versus baseline before the start of the treatment, we used a linear mixed model implemented using the R package nlme:

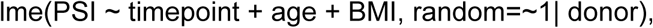

where the timepoint is either 0 months (baseline) or 6 months, and age and BMI are the (scaled) values measured at the corresponding timepoint. To further assess the PSI change independent of sex hormone levels, we fitted three additional models with additional covariates i) testosterone, ii) estradiol, and iii) testosterone and estradiol, all measured both at 0 months and at 6 months. The two treatment arms (CPA and SPIRO) were analyzed separately, and the results were then meta-analyzed using the R package metafor (version 4.8).

Within the FinnGen cohort, we generated proteomic data from individuals with evidence of masculinizing gender affirming treatment (N=23). The inclusion criteria were as follows: 1) at least one occurrence of ICD-10 code F64.0 (“transgenderism”) in the inpatient or outpatient health register data; 2) at least one testosterone purchase in the drug purchase register; and 3) sample date after the first testosterone purchase. All individuals in this subset have XX chromosomes, and proteomic data were measured using the Olink Explore HT panel. We used FinnGen blood donors (N=1,413) to normalize the NPX values to 0 mean and 1 standard deviation by training a scaler in the blood donors and applying these scaling factors to all individuals. We computed the PSI values within these individuals with evidence of masculinizing GAHT and all Olink Explore blood donors with XX or XY chromosomes. We then estimated the association between PSI and GAHT by fitting a regression model with F64 status as the outcome and PSI and age as the covariates, using blood donors with XX chromosomes as the controls.

### Associations of PSI with sex chromosome aneuploidies and mosaic loss of X and Y

We tested associations between PSI and sex chromosome trisomies using linear regression models. We used MoChA ^54,55^ to identify individuals with XXY, XYY, and XXX karyotypes in the UK Biobank test set. Individuals with an XY karyotype were used as controls for XXY and XYY comparisons, and individuals with XX as controls for XXX. All models were adjusted for age.

We replicated the association between PSI and XXY within FinnGen, using individuals with Klinefelter syndrome (XXY; N=17) as cases and blood donors (N=1,413) as controls. Within this replication analysis, we used the 28-protein PSI model, due to proteomics in these individuals being measured using the Olink Explore HT panel, and derived PSI within the FinnGen Olink Explore HT blood donors and XXY individuals combined. The association analysis was conducted the same way as in the UK Biobank, using individuals with XY chromosomes as controls. We further repeated the PSI-XXY association using LOPO-PSI values in the UK Biobank to assess whether the association is driven by a single protein. To investigate the relationship between the PSI proteins and XXY, we fit age-adjusted linear regression models using each of the 31 PSI proteins as the predictor and XXY status as the outcome, and compared the effect sizes to each protein’s sex bias.

Mosaic loss of sex chromosomes was similarly identified using MoChA. For mosaic loss of X (mLOX), individuals with XX karyotypes were used as controls; for mosaic loss of Y (mLOY), we used individuals with XY as controls. These associations were tested using linear models adjusted for age, with an additional model including age-squared to account for potential nonlinear effects.

To account for limited sample sizes, we used additional permutation testing to assess statistical significance. For each model, the outcome was randomly shuffled 1,000 times, and empirical p-values were calculated as the proportion of permutations with absolute effect sizes greater than or equal to the observed.

### Associations of PSI with disease incidence

We computed associations between PSI and incidence disease outcomes in the UK Biobank using the Cox proportional hazards model, run separately by chromosomal sex. Disease phenotypes were defined by mapping 4-character ICD-9 and ICD-10 inpatient diagnoses to Phecodes ^35^ using the PheWAS R package (version 0.99.6-1). Individuals with at least one occurrence of a given code were classified as cases, and the date of first occurrence was used as the disease onset date. We focused on top-level (without decimals) Phecodes in the analyses and excluded all Phecodes with less than 20 cases in either chromosomal sex within the UK Biobank test set. Importantly, we use the built-in control exclusions for Phecodes, designed to prevent contamination of controls with individuals with very similar diagnoses to the diagnosis that defines the cases.

The follow-up period was defined as the period starting three months after the initial assessment center visit (date of proteomic measurement; 2006 to 2010) and ending at the first occurrence of the Phecode of interest, death, or the censoring date (May 31, 2022; minimum of the UK Biobank censoring dates), whichever occurred first. The three-month washout period was applied to reduce potential bias from undiagnosed conditions already present at the time of proteomic measurement. Individuals were considered cases if the first diagnosis of the target Phecode occurred during follow-up. Individuals with the target Phecode or any of its exclusion diagnoses recorded prior to the start of follow-up were excluded. All other individuals were used as controls.

Models were fitted using the survival R package (version 3.5-7). For both chromosomal sex groups, we adjusted the models for age (UKB Data-Field 53; ‘Date of attending assessment centre’), body mass index (UKB Data-Field 21001; ‘Body mass index (BMI)’), serum creatinine (UKB Data-Field 30700; ‘Creatinine’), testosterone (UKB Data-Field 30850; ‘Testosterone’), deprivation index (UKB Data-Field 22189; ‘Townsend deprivation index at recruitment’), smoking status (UKB Data-Field 20116; ‘Smoking status’; categorical), and alanine aminotransferase (UKB Data-Field 30620; ‘Alanine aminotransferase’). For females, we additionally adjusted for hormone-replacement therapy (UKB Data-Field 2814; ‘Ever used hormone-replacement therapy’) and menopause status (UKB Data-Field 2724; ‘Had menopause’).

To understand how unique the observed disease associations are to the 31 proteins included in PSI, we calculated the association between NPX of each protein and the incidence of diseases observed to be significantly associated with PSI. For this analysis, the NPX was inverse rank-normalized within the chromosomal sexes similarly to PSI, and the associations were computed using the Cox proportional hazards model using the same covariates as for PSI. To estimate how unique the observed associations are to the selected proteins, we sampled 1,000 sets of 31 proteins with replacement from non-PSI proteins with Pearson correlation <0.1 with any of the 31 PSI proteins (376 proteins). We then compared both the number of unique associations and the mean strength of the associations between these samples and the 31 PSI proteins.

### Associations of PSI with COVID-19 severity

We assessed the association between PSI and COVID-19 severity using data from 1,372 individuals in the JCTF. COVID-19 severity was categorized into three groups: mild/asymptomatic, hospitalized, and critical. Associations between PSI and COVID-19 severity were tested separately in males (XY sex chromosomes) and females (XX sex chromosomes) using linear regression, adjusting for age at admission.

### Assessing the added value of PSI over sex in model fit

We evaluated whether including PSI, modeled with an interaction term with chromosomal sex, improved model fit compared to a base model including only sex. The two models were adjusted for age, BMI, creatinine, testosterone, alanine aminotransferase, Townsend deprivation index, and smoking status, as in the sex-stratified analyses. We also conducted secondary analyses including age as the only additional covariate. We restricted analyses to diseases that were present in both sex groups, defined as having at least 20 male and 20 female cases, and excluded sex-specific diseases based on Phecode definitions (PheWAS catalog, version 1.2).

We assessed the model fit using Akaike information criterion (AIC), likelihood ratio tests (LRT), and the net reclassification index (NRI). For AIC, we compared values between PSI and sex models, where lower values indicate better fit after accounting for model complexity, and considered differences greater than four (|ΔAIC|>4) in either direction as evidence of improved model fit. For LRT, we compared the nested models to test whether the PSI provided a better fit, applying FDR correction across Phecodes. For NRI, we quantified whether including PSI improved individual-level risk prediction by examining reclassification, with Bonferroni-corrected 95 % confidence intervals used to assess significance.

### Twin-based heritability of PSI

We estimated the twin-based heritability of PSI using data from 401 same-sex (according to sex chromosome group) twins with quality-controlled proteomic data ^56^. PSI was computed using the previously trained XGBoost model; CGA, one of the 31 PSI proteins, was not available in this dataset and was imputed as zero. Individuals were excluded if their predicted sex was fully discordant with genetic sex (distance between predicted and chromosomal sex > 0.95), and their co-twins were also removed. After exclusions, the sample consisted of 111 monozygotic and 79 dizygotic twin pairs. We estimated intra-pair correlations using linear mixed effects models with family identifiers as a random effect, and estimated the heritability in the combined-sex sample using Falconer’s formula.

### Genome-wide association studies of PSI

We conducted two genome-wide association studies (GWAS) of PSI, one within males (XY sex chromosomes) and another one within females (XX sex chromosomes), using PSI as a quantitative phenotype. We restricted the sample to those with European genetic ancestry. Association testing was conducted using linear regression models implemented in Regenie (version 3.4), regressing PSI on genotypes. All models were adjusted for age and 20 genetic principal components. Genome-wide significance was defined as p < 5×10^−8^.

To estimate autosomal SNP-based heritability of PSI, we applied High-Definition Likelihood ^57^ software using GWAS summary statistics from the sex-stratified analyses.

For each chromosomal sex, we identified independent genome-wide significant loci and used LDlink ^58^ to assess whether the lead variants were in linkage disequilibrium across sexes. We annotated the top variants using cis-pQTL data from the FinnGen Green Sandbox, focusing on the most significant associations within the 31 PSI proteins. We further evaluated overlap with sex-biased cis-pQTLs (sb-pQTL) ^14^, focusing on whether PSI-associated loci correspond to pQTLs with differential effects by sex. Disease associations for the top variants were obtained from the FinnGen Results Browser (results.finngen.fi, release 12).

To test whether the PSI proteins were enriched for sex-biased pQTLs (sb-pQTL) compared to other proteins, we tested for overlap with the list of sex-differential pQTLs reported by Koprulu et al. ^14^. We performed a one-sided hypergeometric test to compute the number of overlapping proteins exceeding what would be expected by chance, using the set of all 2,919 Olink-measured proteins as the background.

### PSI polygenic score (PGS)

The PGS SNP weights of PSI were calculated using LDpred2-auto ^59^, implemented in the R package bigsnpr with default settings. We used the HapMap3+ linkage disequilibrium (LD) matrix provided by the R package to achieve higher genome coverage. After excluding SNPs with minor allele frequency (MAF) below 1 %, we calculated PGS SNP weights for 1,327,234 SNPs in females and 1,327,255 SNPs in males for which LD information was available. We used PLINK2 ^60^ to calculate the PGS in three different cohorts: UK Biobank, FinnGen, and MESA. The PGS were not calculated for UK Biobank participants included in the PSI GWAS. PSI was regressed on the PGS and the score combining genetic effects from lead variants to validate the predictive power of PGS and lead variants of loci in all three cohorts. The associations between continuous anthropometric and blood assay traits were estimated by linear regression. We used the Cox proportional hazards model to predict disease risk using PGS in both UK Biobank and FinnGen. For all regression analyses, PGS was scaled and models were adjusted for age and 10 genetic principal components.

## Data Availability

UK Biobank data are available through a procedure described at: https://www.ukbiobank.ac.uk/enable-your-research.
Multi-Ethnic Study of Atherosclerosis (MESA) data access procedure is described at: https://internal.mesa-nhlbi.org/about/publications.
FinnGen data can be accessed through the Fingenious services (https://site.fingenious.fi/en/) managed by FINBB.
Biobank Japan application process for obtaining samples and data is available at: https://biobankjp.org/en/researchers/1974.
Proteomic data from the feminizing gender affirming hormone therapy cohort is available at https://www.ebi.ac.uk/pride/archive/affinity-proteomics/PAD000016.

## Acknowledgements

We gratefully acknowledge all participants and research teams involved in UK Biobank, FinnGen, MESA, Biobank Japan, the Japan COVID-19 Task Force, and Finnish Twin Cohort for making this study possible.

Whole genome sequencing (WGS) for the Trans-Omics in Precision Medicine (TOPMed) program was supported by the National Heart, Lung and Blood Institute (NHLBI). WGS for “NHLBI TOPMed: Multi-Ethnic Study of Atherosclerosis (MESA)” (phs001416.v1.p1) was performed at the Broad Institute of MIT and Harvard (3U54HG003067-13S1). Centralized read mapping and genotype calling, along with variant quality metrics and filtering were provided by the TOPMed Informatics Research Center (3R01HL-117626-02S1). Phenotype harmonization, data management, sample-identity QC, and general study coordination, were provided by the TOPMed Data Coordinating Center (3R01HL-120393-02S1). The MESA projects are conducted and supported by the National Heart, Lung, and Blood Institute (NHLBI) in collaboration with MESA investigators. Support for MESA is provided by contracts 75N92025D00022, 75N92020D00001, HHSN268201500003I, N01-HC-95159, 75N92025D00026, 75N92020D00005, N01-HC-95160, 75N92020D00002, N01-HC-95161, 75N92025D00024, 75N92020D00003, N01-HC-95162, 75N92025D00027, 75N92020D00006, N01-HC-95163, 75N92025D00025, 75N92020D00004, N01-HC-95164, 75N92025D00028, 75N92020D00007, N01-HC-95165, N01-HC-95166, N01-HC-95167, N01-HC-95168, N01-HC-95169, UL1-TR-000040, UL1-TR-001079, UL1-TR-001420, UL1TR001881, and R01HL105756. The authors thank the MESA participants and the MESA investigators and staff for their valuable contributions. A full list of participating MESA investigators and institutions can be found at http://www.mesa-nhlbi.org.

We thank M. Huuska and the Gender Diversity & Intersex Centre of Expertise (SMOK) for guidance on communication. We also thank L. Almeida-Souza, A.M. Berner, C. Orkin, C. Pasin, M. Koprulu, T.C. Oakes-Monger, J. Perry, and P. Vartiainen for valuable feedback on the manuscript. We wish to acknowledge CSC – IT Center for Science, Finland, for computational resources.

## Author contributions

Conceptualization: EV, TH, TN, XS, AG

Methodology: EV, TH, MZ, ZY, TN, MF, XS, AG

Software: EV, TH, MZ, ZY, RE, TN, ALa, MK, DF, CCa, KZ, MAA, GD, LE, NNLN

Validation: EV, TH, MZ

Formal analysis: EV, TH, MZ, ZY, TN, GD, NNLN

Investigation: EV, TH, MZ, ZY, RE, TN, TC, CCa, KZ, GD

Resources: FinnGen, Biobank Japan, Japan COVID-19 Task Force, JH, JP, MO, JK, HN, KF, KM, JIR, SSR, KDT, REG, CCl, PYL, ACW, CJ, TS, MD, YO, XS, BN, ASC

Data curation: EV, TH, MZ, ZY, RE, TN, ALa, ALi, MK, SMW, NNLN, BN, ASC

Writing – original draft: EV, TH, XS, AG

Writing – review & editing: EV, TH, MZ, ZY, RE, TN, ALa, MK, NNLN, TC, DF, CCa, KZ, SMW, MAA, GD, LE, MF, RR, JH, JP, MO, JK, HN, KF, MK, JIR, SSR, KDT, REG, CCl, PYL, ACW, CJ, ASC, TS, MD, TT, BN, YO, XS, AG

Visualizations: EV, TH

Supervision: XS, AG

Project administration: EV, TH, AG

Funding acquisition: XS, AG

## Ethics declarations

This research has been conducted using the UK Biobank Resource under Application Number 170315. This work uses data provided by patients and collected by the NHS as part of their care and support.

Study subjects in FinnGen provided informed consent for biobank research, based on the Finnish Biobank Act. Alternatively, separate research cohorts, collected prior the Finnish Biobank Act came into effect (in September 2013) and start of FinnGen (August 2017), were collected based on study-specific consents and later transferred to the Finnish biobanks after approval by Fimea (Finnish Medicines Agency), the National Supervisory Authority for Welfare and Health. Recruitment protocols followed the biobank protocols approved by Fimea. The Coordinating Ethics Committee of the Hospital District of Helsinki and Uusimaa (HUS) statement number for the FinnGen study is Nr HUS/990/2017. The FinnGen study is approved by Finnish Institute for Health and Welfare (permit numbers: THL/2031/6.02.00/2017, THL/1101/5.05.00/2017, THL/341/6.02.00/2018, THL/2222/6.02.00/2018, THL/283/6.02.00/2019, THL/1721/5.05.00/2019 and THL/1524/5.05.00/2020), Digital and population data service agency (permit numbers: VRK43431/2017-3, VRK/6909/2018-3, VRK/4415/2019-3), the Social Insurance Institution (permit numbers: KELA 58/522/2017, KELA 131/522/2018, KELA 70/522/2019, KELA 98/522/2019, KELA 134/522/2019, KELA 138/522/2019, KELA 2/522/2020, KELA 16/522/2020), Findata permit numbers THL/2364/14.02/2020, THL/4055/14.06.00/2020, THL/3433/14.06.00/2020, THL/4432/14.06/2020, THL/5189/14.06/2020, THL/5894/14.06.00/2020, THL/6619/14.06.00/2020, THL/209/14.06.00/2021, THL/688/14.06.00/2021, THL/1284/14.06.00/2021, THL/1965/14.06.00/2021, THL/5546/14.02.00/2020, THL/2658/14.06.00/2021, THL/4235/14.06.00/2021, Statistics Finland (permit numbers: TK-53-1041-17 and TK/143/07.03.00/2020 (earlier TK-53-90-20) TK/1735/07.03.00/2021, TK/3112/07.03.00/2021) and Finnish Registry for Kidney Diseases permission/extract from the meeting minutes on 4th July 2019. The Biobank Access Decisions for FinnGen samples and data utilized in FinnGen Data Freeze 12 include: THL Biobank BB2017_55, BB2017_111, BB2018_19, BB_2018_34, BB_2018_67, BB2018_71, BB2019_7, BB2019_8, BB2019_26, BB2020_1, BB2021_65, Finnish Red Cross Blood Service Biobank 7.12.2017, Helsinki Biobank HUS/359/2017, HUS/248/2020, HUS/430/2021 §28, §29, HUS/150/2022 §12, §13, §14, §15, §16, §17, §18, §23, §58, §59, HUS/128/2023 §18, Auria Biobank AB17-5154 and amendment #1 (August 17 2020) and amendments BB_2021-0140, BB_2021-0156 (August 26 2021, Feb 2 2022), BB_2021-0169, BB_2021-0179, BB_2021-0161, AB20-5926 and amendment #1 (April 23 2020) and it’s modifications (Sep 22 2021), BB_2022-0262, BB_2022-0256, Biobank Borealis of Northern Finland_2017_1013, 2021_5010, 2021_5010 Amendment, 2021_5018, 2021_5018 Amendment, 2021_5015, 2021_5015 Amendment, 2021_5015 Amendment_2, 2021_5023, 2021_5023 Amendment, 2021_5023 Amendment_2, 2021_5017, 2021_5017 Amendment, 2022_6001, 2022_6001 Amendment, 2022_6006 Amendment, 2022_6006 Amendment, 2022_6006 Amendment_2, BB22-0067, 2022_0262, 2022_0262 Amendment, Biobank of Eastern Finland 1186/2018 and amendment 22§/2020, 53§/2021, 13§/2022, 14§/2022, 15§/2022, 27§/2022, 28§/2022, 29§/2022, 33§/2022, 35§/2022, 36§/2022, 37§/2022, 39§/2022, 7§/2023, 32§/2023, 33§/2023, 34§/2023, 35§/2023, 36§/2023, 37§/2023, 38§/2023, 39§/2023, 40§/2023, 41§/2023, Finnish Clinical Biobank Tampere MH0004 and amendments (21.02.2020 & 06.10.2020), BB2021-0140 8§/2021, 9§/2021, §9/2022, §10/2022, §12/2022, 13§/2022, §20/2022, §21/2022, §22/2022, §23/2022, 28§/2022, 29§/2022, 30§/2022, 31§/2022, 32§/2022, 38§/2022, 40§/2022, 42§/2022, 1§/2023, Central Finland Biobank 1-2017, BB_2021-0161, BB_2021-0169, BB_2021-0179, BB_2021-0170, BB_2022-0256, BB_2022-0262, BB22-0067, Decision allowing to continue data processing until 31st Aug 2024 for projects: BB_2021-0179, BB22-0067, BB_2022-0262, BB_2021-0170, BB_2021-0164, BB_2021-0161, and BB_2021-0169, and Terveystalo Biobank STB 2018001 and amendment 25th Aug 2020, Finnish Hematological Registry and Clinical Biobank decision 18th June 2021, Arctic biobank P0844: ARC_2021_1001.

The MESA study was supported by contracts 75N92020D00001, HHSN268201500003I, N01-HC-95159, 75N92020D00005, N01-HC-95160, 75N92020D00002, N01-HC-95161, 75N92020D00003, N01-HC-95162, 75N92020D00006, N01-HC-95163, 75N92020D00004, N01-HC-95164, 75N92020D00007, N01-HC-95165, N01-HC-95166, N01-HC-95167, N01-HC-95168, and N01-HC-95169 from the National Heart, Lung, and Blood Institute and by grants UL1-TR-000040, UL1-TR-001079, and UL1-TR-001420 from the National Center for Advancing Translational Sciences (NCATS). We also thank the other investigators, the staff, and the participants of MESA (Multi-Ethnic Study of Atherosclerosis) for their valuable contributions. A full list of participating MESA investigators and institutions can be found at http://www.mesa-nhlbi.org.

Molecular profiling for the Australian feminizing GAHT study was approved by the Austin Health Human Research Ethics Committee (Ethics Approval Number: HREC/44503/Austin-2018).

## Code availability

The code will be made available on GitHub (https://github.com/dsgelab/proteomic-sex-index) upon publication.

