## Supplementary material for "Sex-related signatures in the plasma proteome not attributable to XX and XY chromosomes and their impact on health and disease"

#### Supplementary Figures

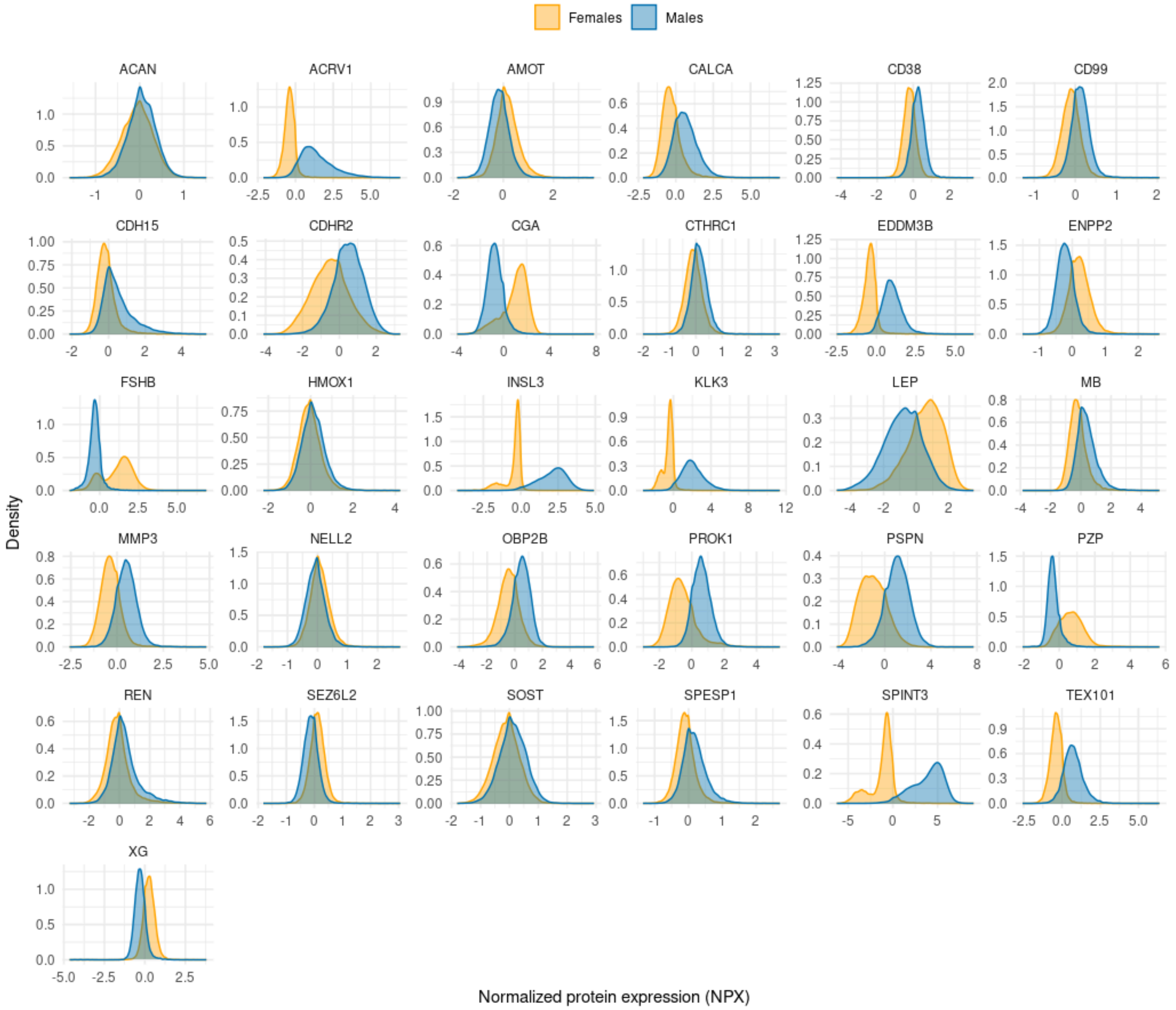


**Supplementary Fig. 1 | Sex-stratified distributions of proteins included in the Proteomic Sex Index (PSI).** Normalized Protein Expression (NPX) values for PSI proteins, shown separately for males (blue) and females (orange). Despite differences between sexes, substantial within-sex variability and between-sex overlap are observed across the proteins.

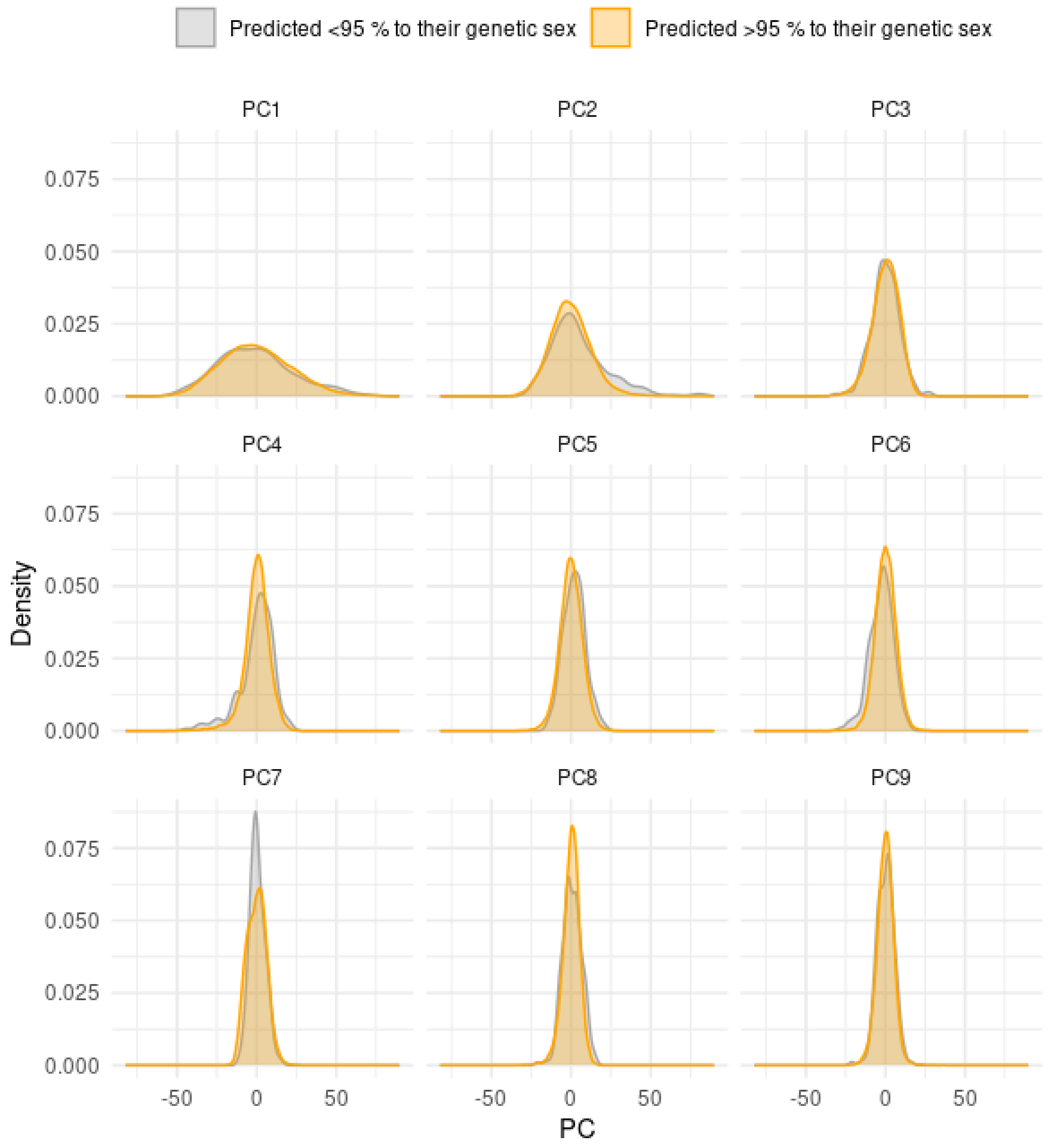


**Supplementary Fig. 2 | Distributions of proteomic principal components by PSI group**. The first nine principal components computed from 2,919 proteins in the UK Biobank, shown for individuals with high (predicted >95% to their genetic sex; orange) and low (predicted <95% to their genetic sex; gray) PSI values. The PSI groups can't be separated based on the first principal components.

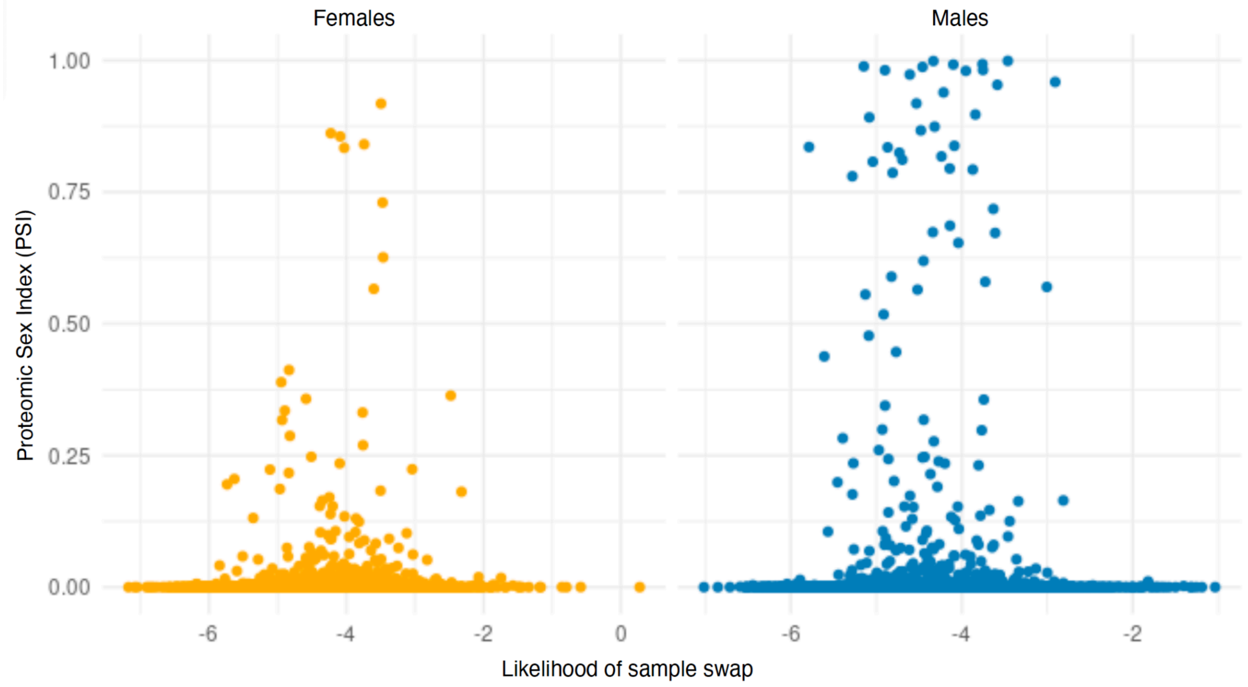


**Supplementary Fig. 3 | Distribution of sample swap likelihood estimates across PSI values.** Relationship between the PSI and sample swap likelihood (scaled log-likelihood ratio) as estimated by Idèfix. Higher values on the x-axis indicate a greater likelihood of sample mix-up.

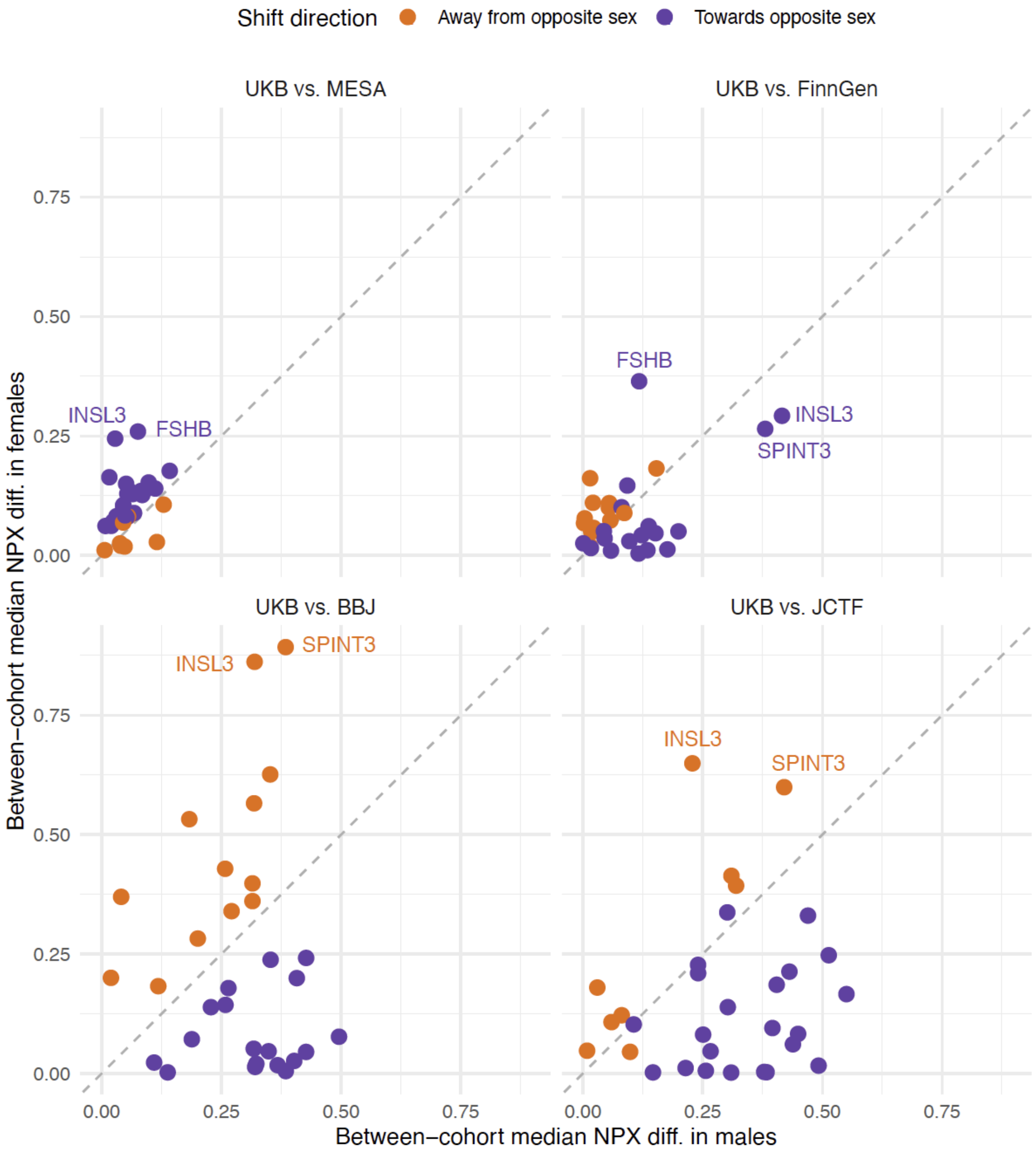


**Supplementary Fig. 4 | Sex-differential shifts in median protein expression across cohorts.** Median NPX values for the 31 PSI proteins in replication cohorts (BBJ, JCTF, MESA, FinnGen), computed separately for males and females, and compared to the corresponding sex-specific medians in the UK Biobank training set, in absolute values. Each protein is annotated by the sex showing the larger absolute shift from the UKB reference, with direction either toward (purple) or away from (red) the opposite sex. Shifts toward the opposite-sex UKB median may lead to increased misclassification in that sex, while shifts away may reinforce sex-typical classification.

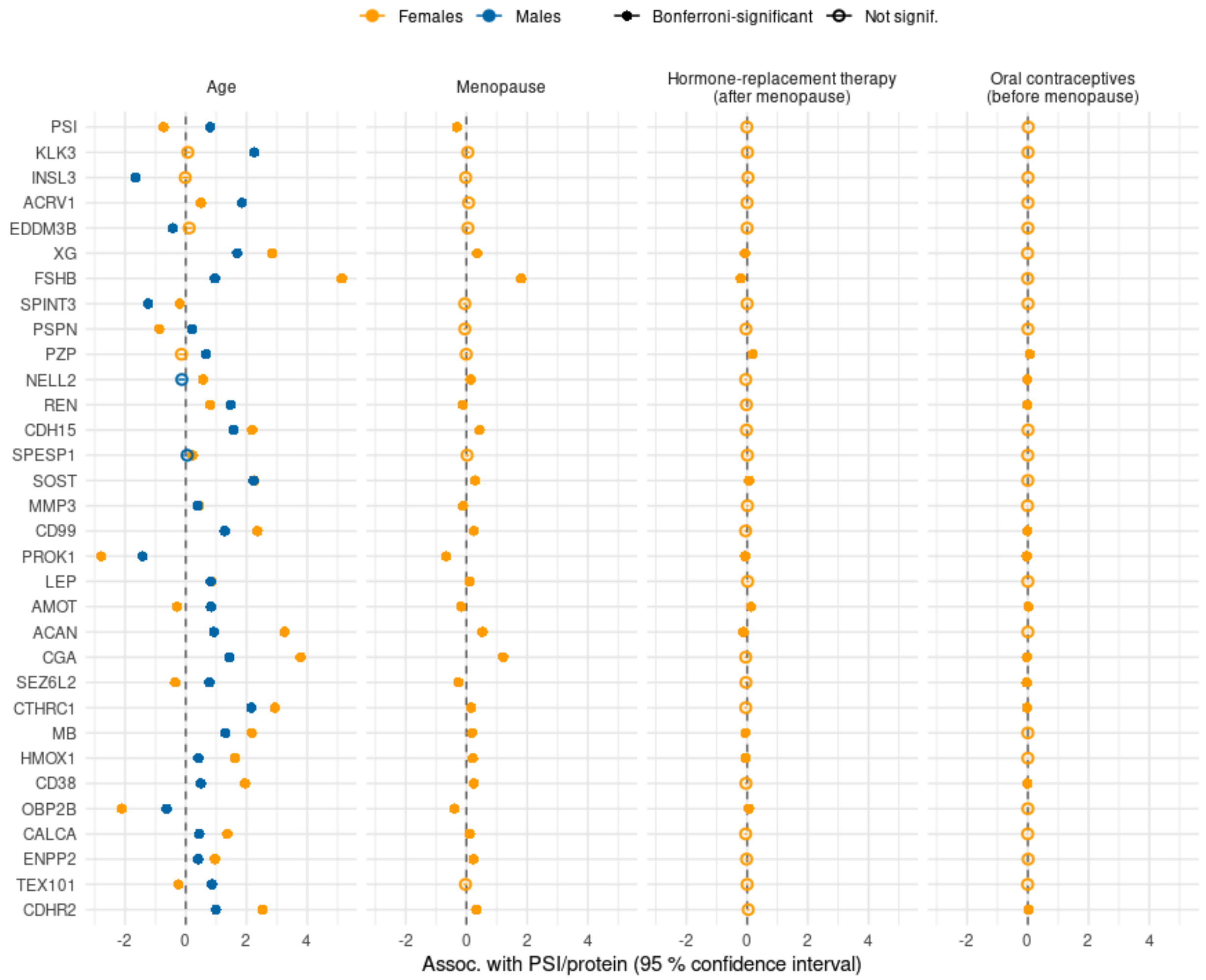


**Supplementary Fig. 5 | Associations of PSI and PSI proteins with age, menopause status, hormone-replacement therapy (HRT), and oral contraceptive use.** Points denote regression coefficients from linear models and error bars indicate 95% confidence intervals. Models were fit separately by genetic sex, and the models addressing the effect of menopause, HRT, and oral contraceptive use are adjusted by age. We included only females who reported having already experienced menopause in the HRT analysis and females before menopause in the oral contraceptive use analysis. PSI is significantly associated with age and menopause status but not with current HRT or oral contraceptive use, despite some PSI proteins showing associations with both.

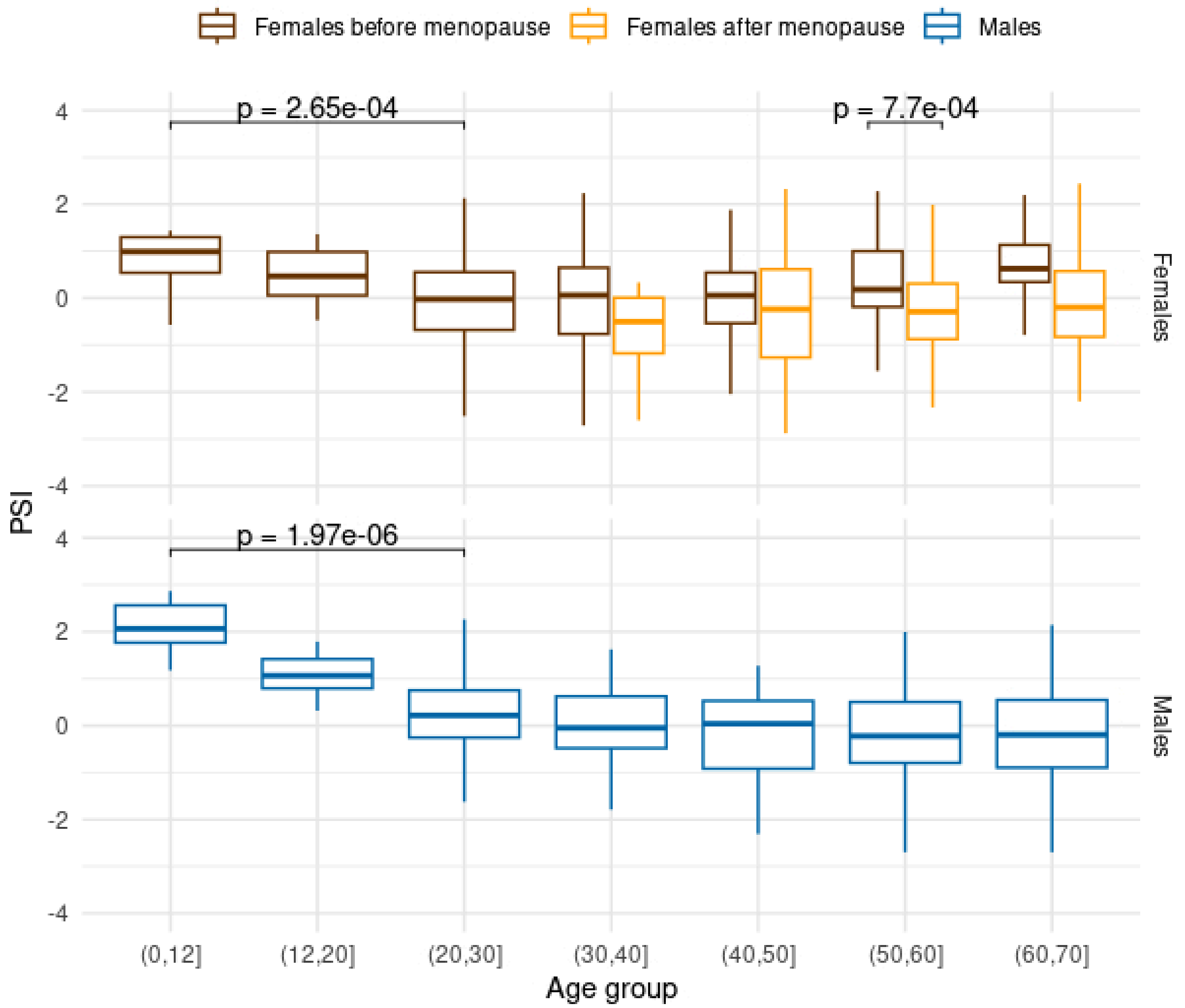


**Supplementary Fig. 6 | Distribution of PSI by age bin and FSHB-derived menopause status in FinnGen (N=1,330).** Boxes denote the range of PSI values in each age bin. Females are grouped by menopausal status based on their follicle-stimulating hormone beta (FSHB) values (see Methods), given that FinnGen does not include information on menopausal status. In short, given that *FSHB* is sensitive to menopausal transition, we see whether their *FSHB* corresponds to those typical of females before or after menopause. Protein expression was measured using Olink Explore HT in all individuals, and we used the 28-protein PSI model to derive PSI. P-values denote unadjusted p-values from the Wilcox test, comparing age groups 0-12 and 20-30 in both sex groups, and the *FSHB*-estimated menopause status within age group 50-60 in females.

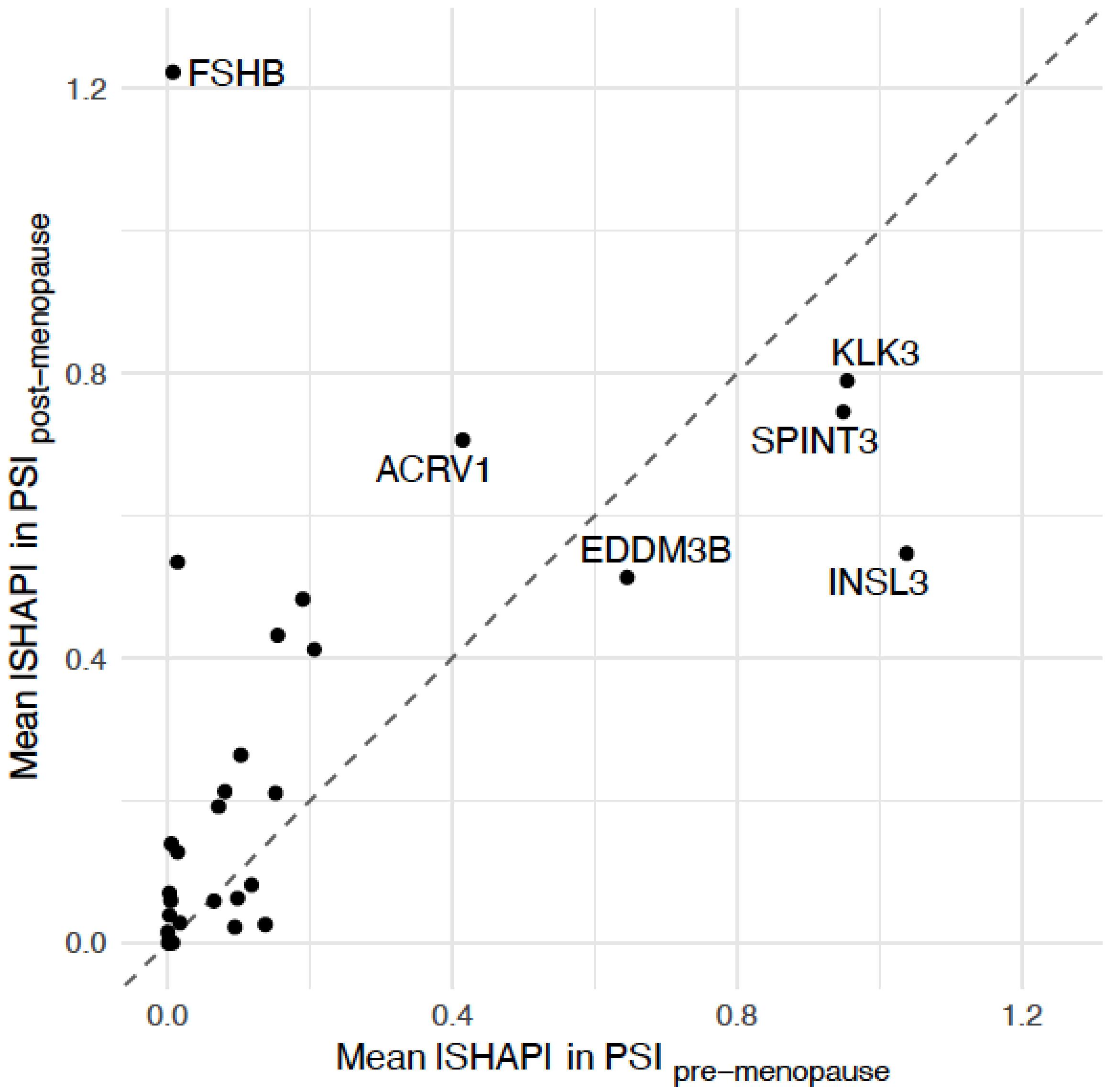


**Supplementary Fig. 7 | Protein importance in menopause-specific PSI models.** Mean absolute SHAP values for proteins in $PSI_{pre\text{-}menopause}$ and $PSI_{post\text{-}menopause}$.

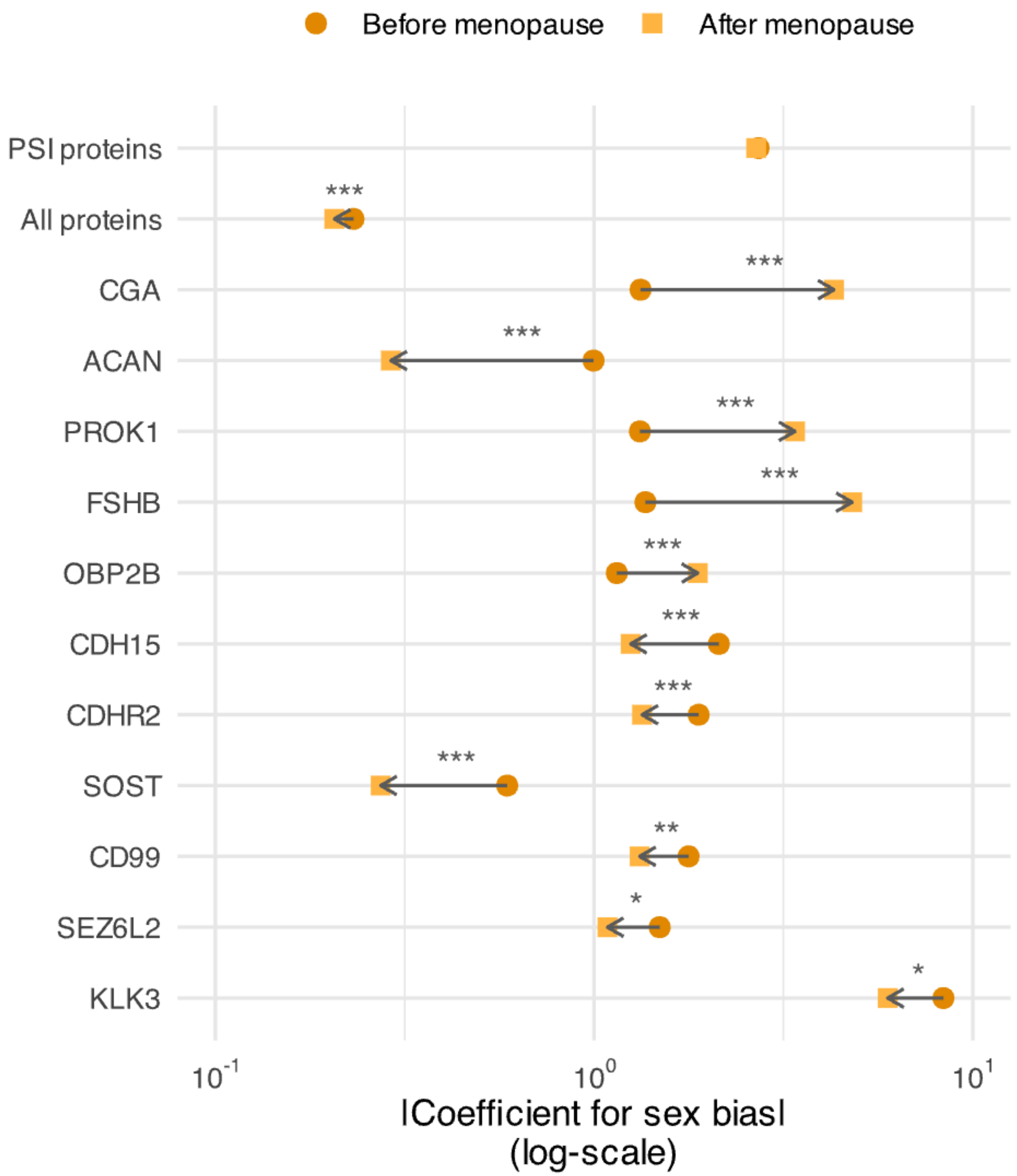


**Supplementary Fig. 8 | Sex-bias before and after menopause relative to age-matched men, among individuals aged 45-55 at the time of recruitment.**

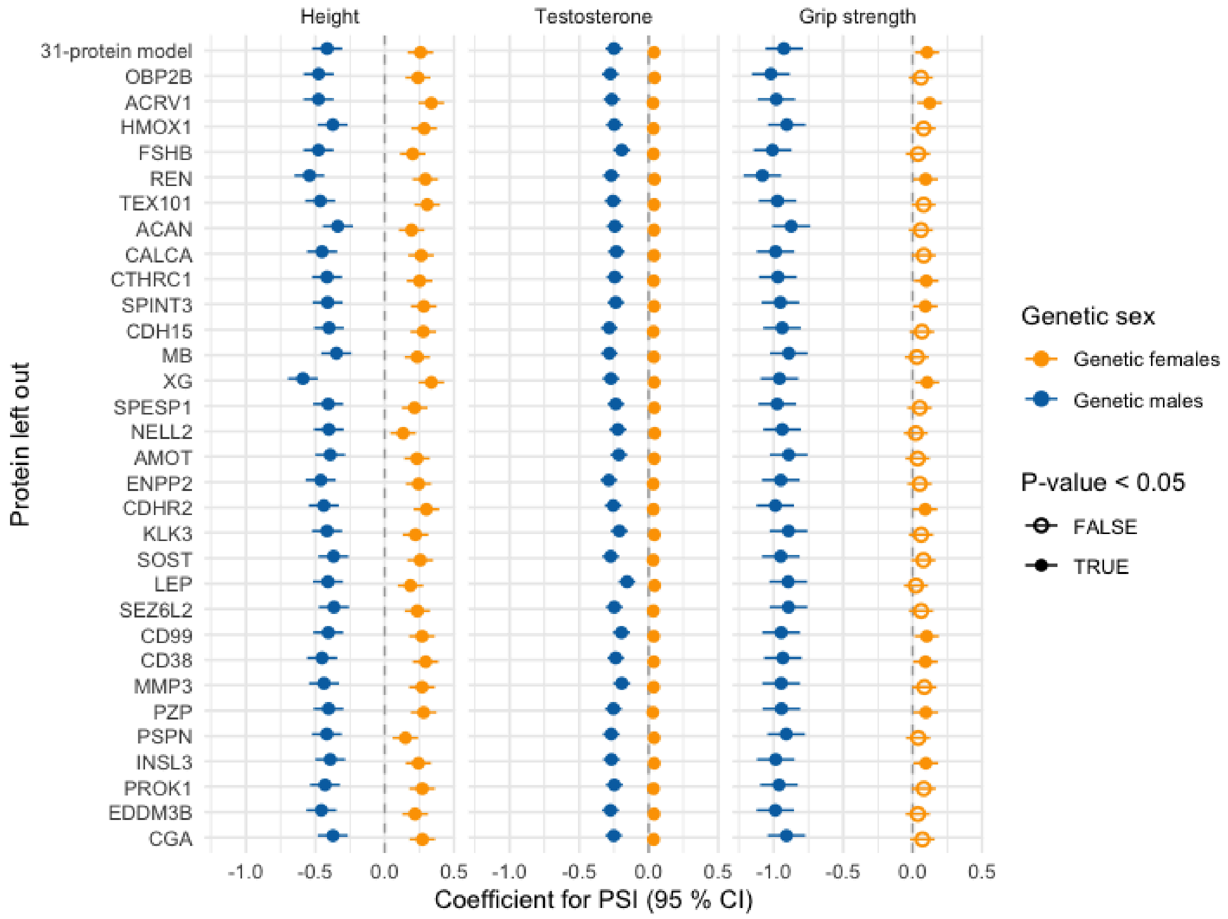


**Supplementary Fig. 9 | Sex-differential phenotype associations for leave-one-protein-out (LOPO) PSI models.**

Associations of LOPO-PSI models with height, testosterone, and grip strength in males and females.

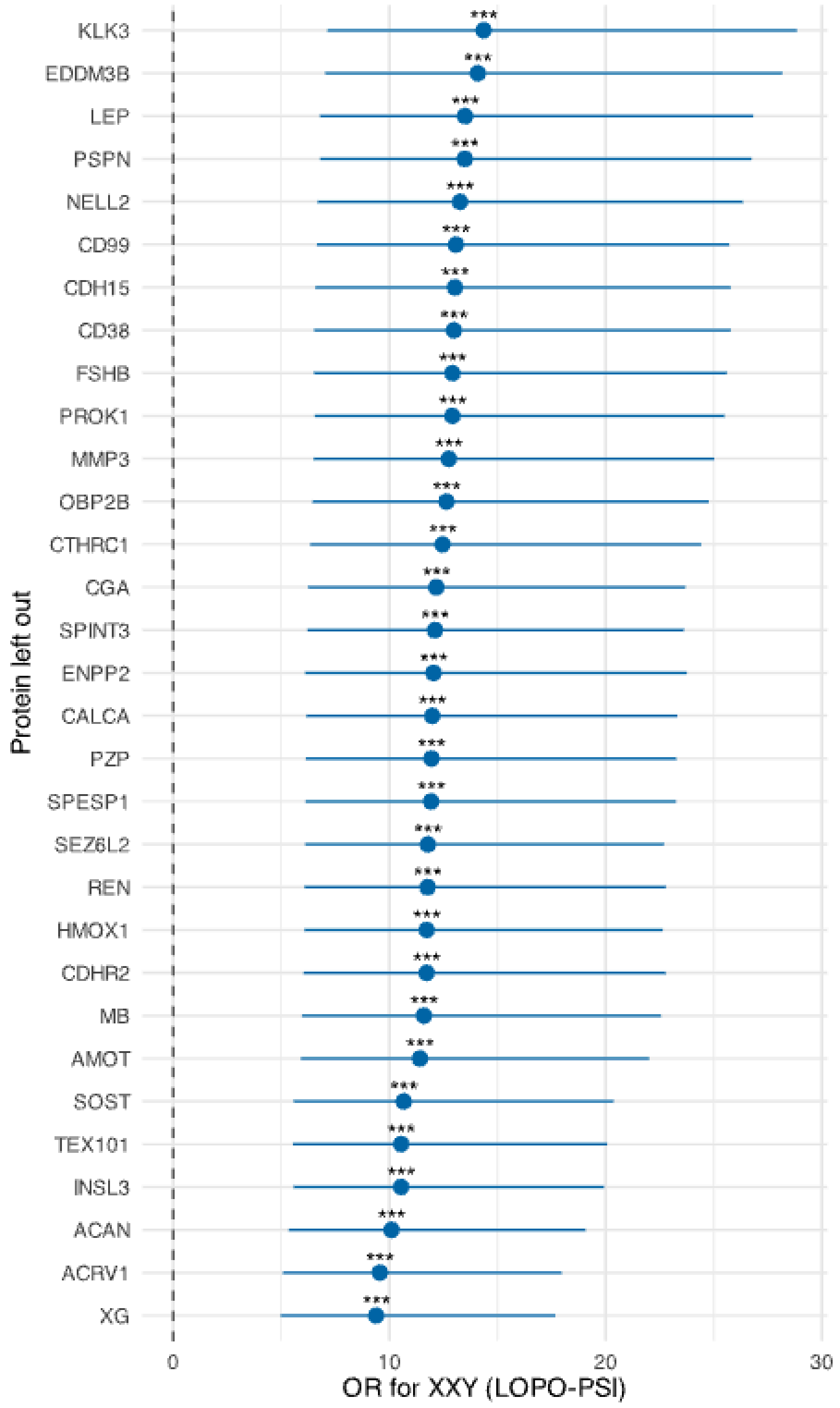


**Supplementary Fig. 10 | Association between LOPO-based PSI models and Klinefelter syndrome (XXY).** Odds ratios (OR) for leave-one-protein-out (LOPO) PSI models with XXY status. Stars (***) indicate Bonferroni-corrected p-value < 0.05.

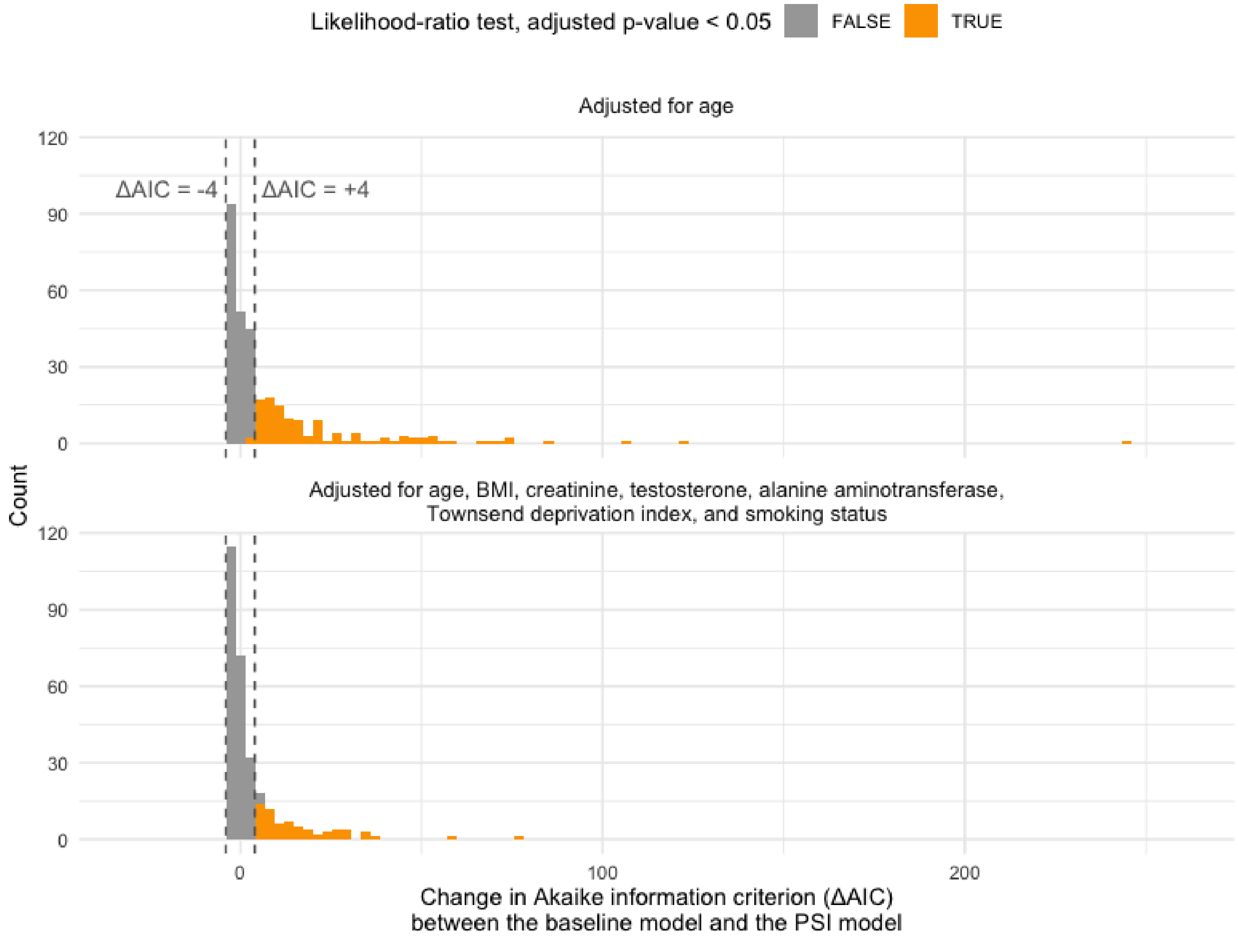


**Supplementary Fig. 11 | Distribution of the differences in Akaike information criterion (ΔAIC) when including the interaction between sex and PSI as a covariate.** ΔAIC values <–4 favor the sex-only model, whereas values >4 favor the model including PSI, after accounting for the increased model complexity.

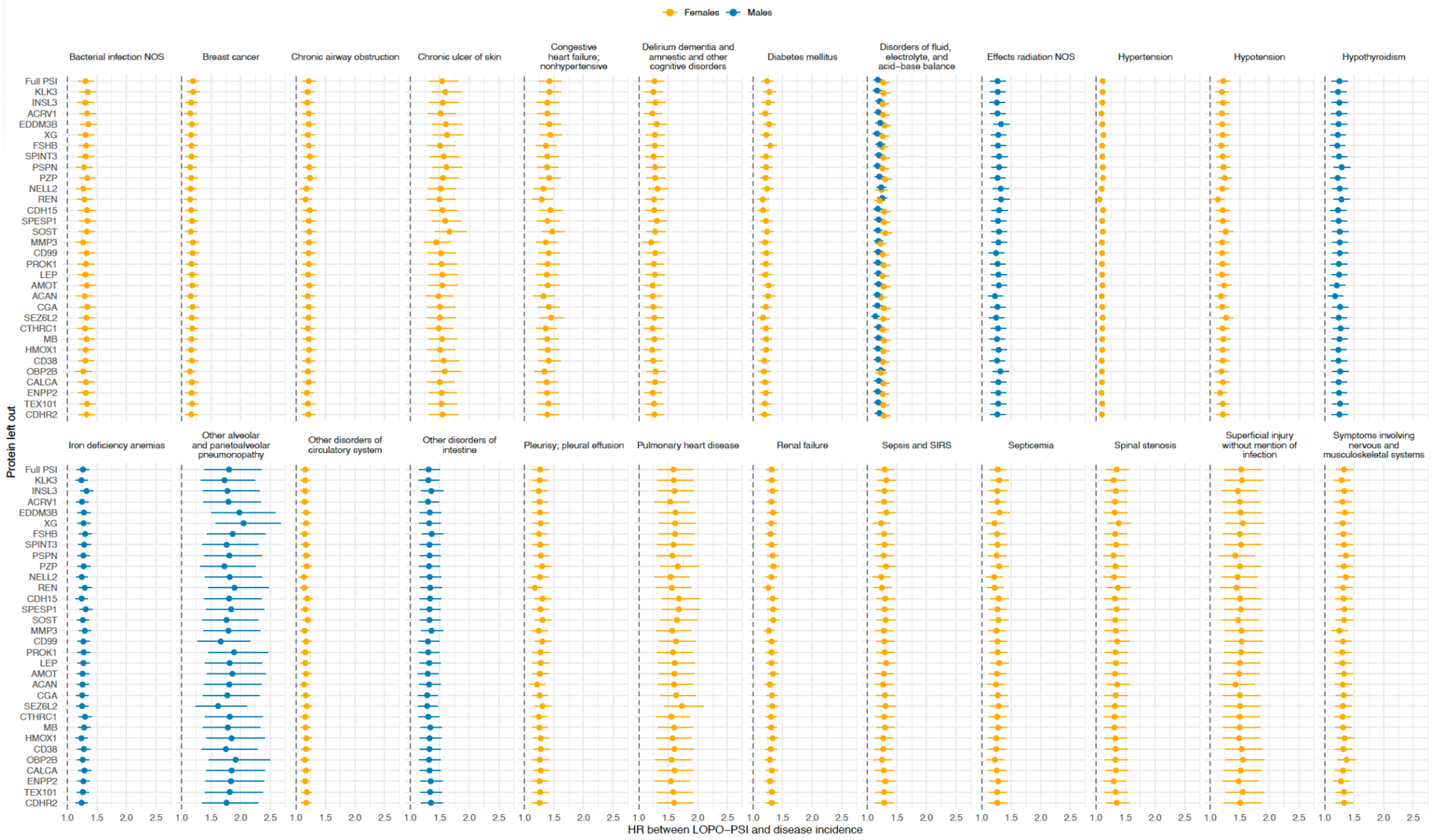


**Supplementary Fig. 12 | Associations between LOPO-based PSI models and disease incidence.** Hazard ratios (HR) from Cox regressions assessing PSI associations with 25 diseases, using leave-one-protein-out (LOPO) models.

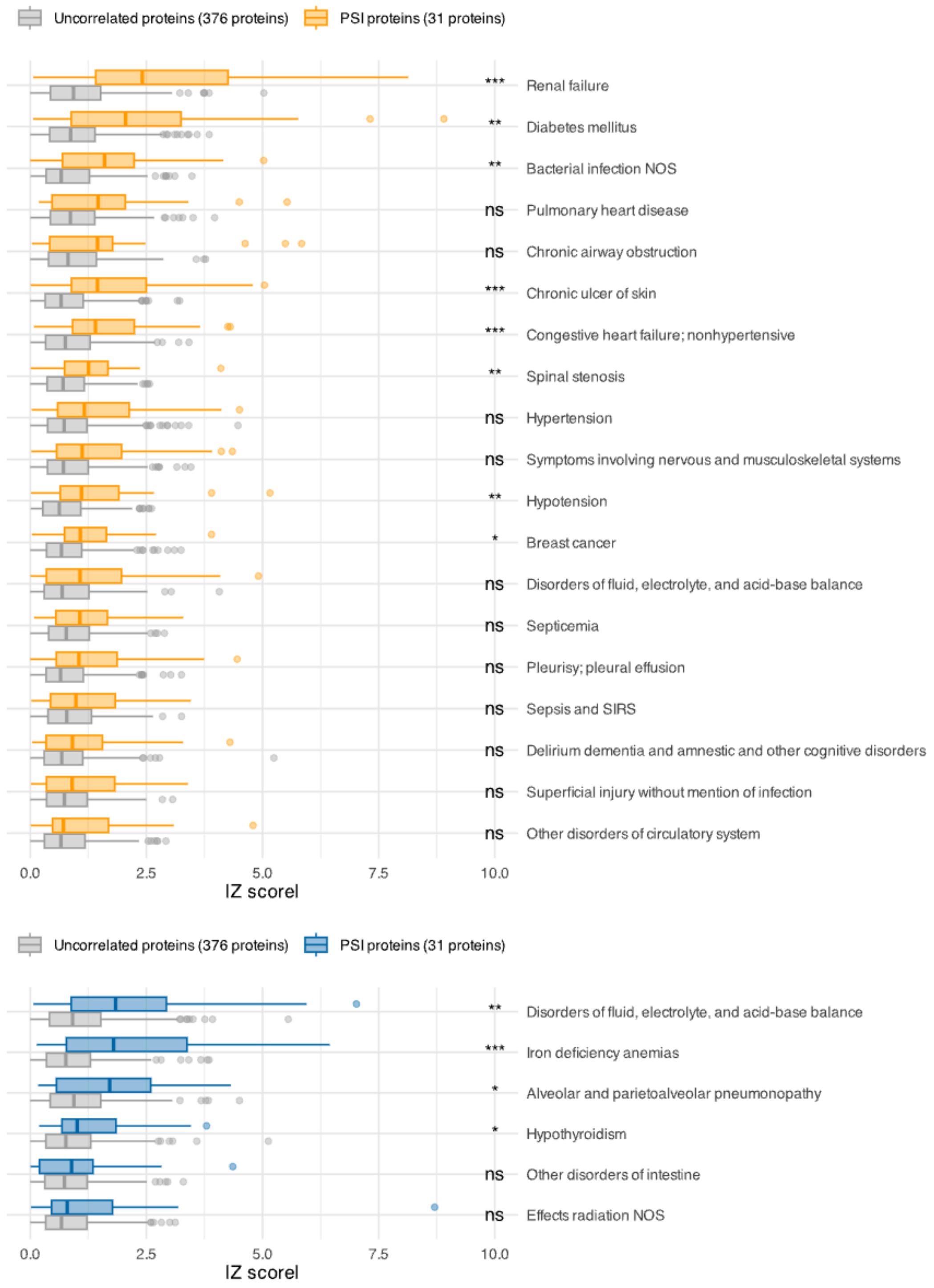


**Supplementary Fig. 13 | Protein-disease associations across all diseases tested.** Absolute Z scores (scaled effect sizes) for associations between 31 PSI proteins (orange for females, blue for males) and 376 other proteins not correlated with PSI proteins (gray; Pearson r < 0.1). Stars indicate significance from Bonferroni-corrected one-sided Wilcoxon tests comparing PSI vs. non-PSI proteins within each disease (***, p < 0.001; **, p < 0.01; *, p < 0.05).

### General audience summary

#### Overview

In this project, we wanted to understand how *chromosomal sex*, whether someone has XX or XY sex chromosomes, can be seen in the proteins in our blood. To explore this, we created the **Proteomic Sex Index (PSI)**, which measures how much a person's blood protein profile resembles typical patterns seen in people with XX or XY chromosomes. PSI offers a measure of sex-related signatures in the proteome, recognizing that some individuals, despite their chromosomal sex, may exhibit a proteomic signature more closely aligned with profiles typical for individuals with different sex chromosomes.

We found that PSI is linked to traits with known differences between chromosomal sexes, such as height, testosterone levels, and grip strength, within each sex. Within individuals receiving gender-affirming hormone treatment, PSI increased after treatment, indicating that it is responsive to hormonal changes. PSI was also connected to risks for certain diseases and differences in immune responses, including COVID-19 severity. We further saw that these patterns are shared by both genetics and the environment.

> *Our study focuses on chromosomal sex and not gender identity. We recognize that gender is important in understanding health and that it exists independently of genetics. Even though we do not have information about an individual's gender, most individuals included in the study, apart from the gender-affirming hormone therapy cohort, are likely cisgender.*
> *We want to demonstrate that acknowledging sex-related signatures as a continuum can help us understand trait expression and disease risk more fully than the binary sex model. We are committed to communicating this research carefully and respectfully.*

#### Why we did this study

Sex and gender affect many aspects of health, such as body size, immune system function, and risk of certain diseases. In research, sex is often treated as a binary trait, yet different factors comprise our understanding of sex, i.e., hormones, external and internal anatomy, and genetics. A binary conception of sex can be overly simplistic – people with the same sex characteristics still vary from one another. This study focuses on one area of these differences: sex chromosomes.

One way to study these differences is by looking at proteins in the blood. Proteins carry out many important roles in the body, such as supporting the immune system, carrying hormones, and sending signals between cells. They are influenced by both genetics and the environment and can show patterns linked to health.

Understanding this variation is important because it may help explain health differences that go beyond simple binary male-female categories. We're especially interested in understanding what it means when someone's blood protein profile resembles patterns more typical of individuals with different sex chromosomes. We hope that insights from this research will help us study health and disease in a way that respects the diversity of human biology.

#### The data we used

For this study, we used blood protein and health data of more than 50,000 individuals from six research cohorts. These datasets include diverse populations from different countries and were collected with informed consent from volunteer participants.

- **UK Biobank** – The UK Biobank is a population-based study of around 500,000 adults in the United Kingdom, recruited between 2006 and 2010. Participants provided biological samples, detailed health information, and consented to long-term follow-up for research purposes. The UK Biobank is one of the largest and most widely used health research resources in the world.
- **FinnGen** – FinnGen is a Finnish public-private partnership combining genetic data from Finnish biobanks with national health registry data. The study includes over 500,000 participants and aims to improve understanding of disease mechanisms and support the development of new treatments. All participants have provided broad research consent for the use of their data and samples.
- **Multi-Ethnic Study of Atherosclerosis** – The Multi-Ethnic Study of Atherosclerosis cohort includes over 6,000 adults from several ethnic backgrounds in the United States. It was designed to investigate the development of cardiovascular disease and includes detailed health information, lifestyle factors, and biological samples.
- **Biobank Japan** – Biobank Japan is one of the largest biobanks in Asia, collecting genetic and clinical information from over 200,000 patients across Japan. It aims to advance personalized medicine and improve understanding of how genetic and environmental factors influence disease.
- **Japan COVID-19 Task Force** – The Japan COVID-19 Task Force is a collaborative research effort focused on understanding the genetic and clinical factors affecting COVID-19. It includes biological samples and health data from patients with different levels of COVID-19 severity.
- **Feminizing gender-affirming hormone treatment (GAHT) cohort** – The feminizing GAHT cohort is based on a clinical trial in Australia of transgender and non-binary individuals assigned male at birth who were starting feminizing gender-affirming hormone therapy with estradiol and antiandrogens. Blood samples were collected before and after treatment, and participants provided consent for their samples to be used in future research.

#### How we defined sex

In this study, we determined sex based on individuals' DNA by looking at their sex chromosomes. We refer to those with XX chromosomes as females and those with XY chromosomes as males. We also included individuals with variations in sex chromosomes, such as having an extra X or Y chromosome (XXX, XXY, and XYY), to study how these differences influence protein patterns in the blood. Importantly, our study focuses only on chromosomal sex and does not include information about other sex characteristics or gender identity.

#### What is the Proteomic Sex Index

The Proteomic Sex Index (PSI) quantified how much a person's blood protein profile resembles the average patterns seen in people with XX or XY chromosomes.

High PSI values indicate a greater difference from the typical protein levels seen in a person's chromosomal sex, while lower values mean their protein levels are similar to those of most people sharing the same sex chromosome configuration as them. People with the same chromosomal sex can have different PSI values. For example, some females might have PSI values indicating patterns more typical of males, and *vice versa*.

PSI was developed using machine learning, based on data from approximately 10,000 individuals with XX or XY chromosomes. By combining information from many proteins, it places each person along a spectrum of sex-related protein patterns.

*The Proteomic Sex Index should not in any way be interpreted as implying that a person's identity or experience can be defined by their blood proteins. PSI is not built using information about gender or gendered behavior, meaning that PSI does not only capture biological processes, but also the potential gendered impact of social and environmental circumstances on people's health. Thus, PSI should not be used to make assumptions related to gender or gender expression. We strongly oppose any misuse of this work to support discrimination, exclusion, or stigma.*

#### How we built and tested the PSI model

To build the PSI model, we started by looking at thousands of proteins measured in blood samples. We focused on the ones that showed clear differences, on average, between people with XX and XY chromosomes. We selected 31 proteins that most robustly reflect these differences in protein levels.

We used a machine learning method called XGBoost to combine information from these proteins and create PSI. We trained this model using data from approximately 10,000 individuals in the UK Biobank. To validate the PSI model, we tested it in over 40,000 individuals not used for training the model in the UK Biobank and four other cohorts: FinnGen, MESA,

Biobank Japan, and the Japan COVID-19 Task Force. We did this to verify that PSI gives consistent results across different populations.

#### What we analyzed

We wanted to study how blood protein patterns that are less typical of the average person with the same sex chromosomes relate to health and biology. Specifically, we looked at:

- **Sex-associated traits**. We tested whether PSI was connected to height, testosterone levels, and grip strength, among people of the same chromosomal sex.
- **Gender-affirming hormone therapy**. We examined whether PSI changes in people receiving gender-affirming hormone therapy by comparing protein patterns before and after treatment to understand how altering hormone levels affects the blood proteome.
- **Sex chromosome differences**. We studied how PSI varies in individuals with sex chromosome configurations different from XX and XY, including Klinefelter syndrome (XXY), Turner syndrome (XXX), and XYY syndrome (XYY), as well as the link between PSI and loss of X and Y chromosomes.
- **Disease risk**. We examined whether people with higher or lower PSI have an increased risk for a wide range of diseases in the future.
- **Genetic influences**. We analyzed how genetics affects PSI, including searching for genetic variants linked to PSI, calculating how much PSI is inherited, and looking at how PSI is genetically related to traits and diseases.

#### What we found

- **Most individuals' protein patterns align with their chromosomal sex, but not everyone's, and PSI captures some of this variation**. While, on average, protein levels differ between chromosomal sexes, there's also significant variation within each of these groups. PSI measures how much a person's protein patterns align with those typically seen in people with XX or XY chromosomes, helping us study differences within these categories.
- **PSI is linked to traits that often differ between sexes.** Males and females differ on average in height, testosterone, and grip strength. We found that among males, those with higher PSI (more female-typical protein patterns) tend to be shorter and have lower testosterone and grip strength. Among females, those with higher PSI (more male-typical protein patterns) tend to be taller, with higher testosterone and greater grip strength.
- **PSI changes with gender-affirming hormone therapy**. In people receiving gender-affirming hormone therapy (GAHT), we observed changes in PSI after treatment, suggesting that blood protein patterns may shift over time during medical transition. These changes were not fully accounted for by measured hormone levels alone, pointing to the involvement of additional biological factors beyond sex hormones.

- **PSI varies with sex chromosome differences**. Individuals with XXY chromosomes (Klinefelter syndrome) tend to have higher PSI compared to those with XY chromosomes, showing protein patterns toward those typical of XX individuals.
- **PSI relates to disease risk**. Higher PSI was linked to the risk of certain diseases within each chromosomal sex. For example, females with higher PSI had a higher risk of future breast cancer, while males with higher PSI were more likely to have iron deficiency anemia. PSI was also linked to diseases not traditionally considered sex-specific, suggesting it may reflect broader biological processes.
- **PSI has a genetic basis and is genetically linked to certain diseases**. Some of the genetic factors linked to PSI are also linked to diseases like breast cancer in females. We also found that these genetic links can differ between chromosomal sexes, showing that different genetic factors may shape PSI in each chromosomal sex.

#### Limitations

- **PSI is based only on specific proteins**. Our study used only certain proteins measured in blood, so PSI may not capture all possible proteins or biological processes related to sex differences.
- **Most individuals' protein patterns align with their chromosomal sex**. Most people's PSI values were similar to average patterns for their sex chromosome group, and only a small number showed larger shifts toward patterns usually seen in people with different sex chromosomes. This means that while PSI helps us study subtle differences within each chromosomal sex, strong shifts are relatively rare.
- **PSI is trained on binary chromosomal sex (XX and XY) and so doesn't capture all variations**. Although PSI provides a continuous measure, it was trained using data from individuals with XX or XY chromosomes. It doesn't account for all variations in sex chromosomes or broader aspects of sex and gender beyond genetics.
- **Limited data on changes over time, including during hormone therapy.** While we observed PSI changes in people receiving gender-affirming hormone therapy, our data include only limited time points. Longer-term studies with repeated measurements are needed to better understand how PSI evolves over time and how such changes relate to health outcomes.
- **We do not have information on gender and traditionally gendered behaviors.** The cohorts used in this study did not include information on individuals' gender identity.

#### Who we are

We are a group of scientists from universities and research institutions in Finland, USA, Japan, UK, China, Australia, and Sweden, interested in studying sex-related variability in the proteome.


- **Essi Viippola**: Institute for Molecular Medicine Finland, Helsinki, Finland; Broad Institute of MIT and Harvard, Boston, USA; Massachusetts General Hospital, Boston, USA
- **Tuomo Hartonen**: Institute for Molecular Medicine Finland, Helsinki, Finland
- **Meihui Zhu**: Institute for Molecular Medicine Finland, Helsinki, Finland
- **Zhijian Yang**: Institute for Molecular Medicine Finland, Helsinki, Finland
- **Ryuya Edahiro**: Osaka University, Suita, Japan; RIKEN Center for Integrative Medical Sciences, Yokohama, Japan
- **Tomoko Nakanishi**: Institute for Molecular Medicine Finland, Helsinki, Finland; University of Tokyo, Tokyo, Japan; Queen Mary University of London, London, UK
- **Ao Lan**: Fudan University, Guangzhou, China
- **Masahiro Kanai**: Broad Institute of MIT and Harvard, Boston, USA; Massachusetts General Hospital, Boston, USA
- **Nhi N. L. Nguyen:** Molecular Immunity, Infection, Immunity and Global Health Theme, Murdoch Children's Research Institute, Royal Children's Hospital, Parkville, Victoria, Australia
- **Tatiana Cajuso**: Institute for Molecular Medicine Finland, Helsinki, Finland
- **Daniela Fusco**: Institute for Molecular Medicine Finland, Helsinki, Finland; University of Turin, Turin, Italy
- **Chiara Caime**: Institute for Molecular Medicine Finland, Helsinki, Finland
- **Kristina Zguro**: Institute for Molecular Medicine Finland, Helsinki, Finland
- **Sebastian May-Wilson**: Institute for Molecular Medicine Finland, Helsinki, Finland
- **M. Austin Argentieri**: Broad Institute of MIT and Harvard, Boston, USA; Massachusetts General Hospital, Boston, USA
- **Gabin Drouard**: Institute for Molecular Medicine Finland, Helsinki, Finland
- **Lisa Eick**: Institute for Molecular Medicine Finland, Helsinki, Finland
- **Matteo Ferro**: Institute for Molecular Medicine Finland, Helsinki, Finland
- **Aoxing Liu**: Institute for Molecular Medicine Finland, Helsinki, Finland; Broad Institute of MIT and Harvard, Boston, USA; Massachusetts General Hospital, Boston, USA
- **Rodosthenis Rodosthenous**: Institute for Molecular Medicine Finland, Helsinki, Finland
- **Jarno Honkanen**: Finnish Red Cross Blood Service, Helsinki, Finland
- **Jukka Partanen**: Finnish Red Cross Blood Service, Helsinki, Finland
- **Miina Ollikainen**: Institute for Molecular Medicine Finland, Helsinki, Finland
- **Jaakko Kaprio**: Institute for Molecular Medicine Finland, Helsinki, Finland
- **Ho Namkoong**: Keio University School of Medicine, Tokyo, Japan
- **Koichi Fukunaga**: Keio University School of Medicine, Tokyo, Japan
- **Koichi Matsuda**: University of Tokyo, Tokyo, Japan
- **Jerome I. Rotter**: The Lundquist Institute for Biomedical Innovation at Harbor-UCLA Medical Center, Torrance, CA, USA
- **Stephen S. Rich**: University of Virginia, Charlottesville, VA, USA

- **Kent D. Taylor**: The Lundquist Institute for Biomedical Innovation at Harbor-UCLA Medical Center, Torrance, CA, USA
- **Robert E. Gerszten**: Beth Israel Deaconess Medical Center, Boston, MA, USA
- **Clary Clish**: Broad Institute of MIT and Harvard, Boston, USA; Massachusetts General Hospital, Boston, USA
- **Peter Y. Liu**: The Lundquist Institute, Torrance, CA, USA; Harbor-UCLA Medical Center, Torrance, CA, USA
- **Alexis C. Wood**: Baylor College of Medicine, Houston, TX, USA
- **Craig Johnson**: University of Washington, Seattle, WA, USA
- **Ada S. Cheung:** Department of Medicine (Austin Health), The University of Melbourne, Heidelberg, Victoria, Australia; Department of Endocrinology, Austin Health, Heidelberg, Victoria, Australia
- **Tamar Sofer**: Brigham and Women's Hospital, Boston, MA, USA; Beth Israel Deaconess Medical Center, Boston, MA, USA
- **Mark Daly**: Institute for Molecular Medicine Finland, Helsinki, Finland; Broad Institute of MIT and Harvard, Boston, USA; Massachusetts General Hospital, Boston, USA
- **Taru Tukiainen**: Institute for Molecular Medicine Finland, Helsinki, Finland
- **Boris Novakovic:** Molecular Immunity, Infection, Immunity and Global Health Theme, Murdoch Children's Research Institute, Royal Children's Hospital, Parkville, Victoria, Australia; Department of Paediatrics, The University of Melbourne, Parkville, Victoria, Australia
- **Yukinori Okada**: University of Tokyo, Tokyo, Japan; RIKEN Center for Integrative Medical Sciences, Yokohama, Japan; Osaka University Graduate School of Medicine, Suita, Japan
- **Xia Shen**: Fudan University, Guangzhou, China; Karolinska Institutet, Stockholm, Sweden
- **Andrea Ganna**: Institute for Molecular Medicine Finland, Helsinki, Finland; Broad Institute of MIT and Harvard, Boston, USA; Massachusetts General Hospital, Boston, USA

We are grateful to the **Gender Diversity & Intersex Centre of Expertise** (*Sukupuolen moninaisuuden osaamiskeskus, SMOK*) and the **SHARE** collaborative for their guidance in helping us ensure that our communication is thoughtful, inclusive, and sensitive to the broader social context in which sex and gender are discussed. We are also thankful to several researchers outside our team who shared valuable feedback on how to communicate these findings clearly and respectfully.

If you have any questions or comments about our study, we would be happy to hear from you. Please contact **Andrea Ganna**.
